# Brain and Blood Transcriptome-Wide Association Studies Identify Five Novel Genes Associated with Alzheimer’s Disease

**DOI:** 10.1101/2024.04.17.24305737

**Authors:** Makaela A. Mews, Adam C. Naj, Anthony J. Griswold, Alzheimer’s Disease Genetics Consortium, Jennifer E. Below, William S. Bush

## Abstract

**INTRODUCTION:** Transcriptome-wide Association Studies (TWAS) extend genome-wide association studies (GWAS) by integrating genetically-regulated gene expression models. We performed the most powerful AD-TWAS to date, using summary statistics from *cis*-eQTL meta-analyses and the largest clinically-adjudicated Alzheimer’s Disease (AD) GWAS.

**METHODS:** We implemented the OTTERS TWAS pipeline, leveraging *cis*-eQTL data from cortical brain tissue (MetaBrain; N=2,683) and blood (eQTLGen; N=31,684) to predict gene expression, then applied these models to AD-GWAS data (Cases=21,982; Controls=44,944).

**RESULTS:** We identified and validated five novel gene associations in cortical brain tissue (*PRKAG1*, *C3orf62*, *LYSMD4*, *ZNF439*, *SLC11A2*) and six genes proximal to known AD-related GWAS loci (Blood: *MYBPC3*; Brain: *MTCH2*, *CYB561*, *MADD*, *PSMA5*, *ANXA11*). Further, using causal eQTL fine-mapping, we generated sparse models that retained the strength of the AD-TWAS association for *MTCH2*, *MADD*, *ZNF439*, *CYB561*, and *MYBPC3*.

**DISCUSSION:** Our comprehensive AD-TWAS discovered new gene associations and provided insights into the functional relevance of previously associated variants.

## 1 BACKGROUND

Alzheimer’s disease (AD) has a strong genetic component, with heritability (*h^2^*) estimates ranging from *h^2^*∼60-80% based on twin studies [1,2]. However, known AD risk variants discovered through genome-wide association studies (GWAS) explain only ∼30% of the genetic variance in disease risk while the remaining ∼70% is attributed to undiscovered AD variants [3]. Multiple recent large-scale GWAS [4,5] have been conducted with varying stringency in phenotyping criteria, with some studies using AD-by-proxy (GWAX) phenotypes [6] that may increase study heterogeneity [7] but also dramatically increase sample size. These GWAS have been successful at identifying new AD risk variants, though nearly all of these variants fall in non-coding regions making their roles in disease risk difficult to interpret. A key component to the translation of these findings into drug targets is understanding how these non-coding variants influence gene expression [8–10]. Transcriptomic integration with GWAS has the potential to both improve statistical power for discovery of novel genetic associations and accelerate associated drug development by mapping variants to their functional outcomes.

Mirroring the advancement of GWAS, studies of expression quantitative trait loci (eQTLs) --which were initially small in scale --have now been aggregated via meta-analysis for extremely well-powered studies [11,12]. While eQTLs generally have larger effects and are detectable in smaller sample sizes than variants associated with disease [13], prior work examining the genetic architecture of *cis*-regulatory variation [14] demonstrates that weaker eQTLs form polygenic components that complement strong, sparse eQTL effects [12]. While TWAS methods were originally developed to use individual-level data [15], recent methodological developments have extended these approaches to allow for the use of summary statistics in both the creation of expression prediction models and for association analysis [16–25].

Previous transcriptome-wide association studies (TWAS) for AD have utilized smaller-scale eQTL reference datasets applied to both clinical AD and AD-by-proxy individual-level and summary data. Methodological extensions to the original TWAS framework like BGW-TWAS [26], T-GEN [27], VC-TWAS [28], UTMOST [29,30], InTACT [31], and MR-JTI [32,33] have been developed and applied to AD, leading to improvements in gene discovery by incorporating *trans*-eQTLs, epigenetic annotations, random *cis*-eQTL effect modeling, and multi-tissue modeling [34] that leverages shared tissue expression profiles to boost power. While these studies have advanced our understanding of how heritable gene expression is associated with AD risk, they have identified varying numbers of significant AD TWAS associations, from 8 [35] to 415 genes [32]. However, there has been limited functional validation or fine mapping of the results. Despite methodological improvements, most of these studies have relied on Genotype-Tissue Expression (GTEx) [36] data for model training. While GTEx provides genetic data across 54 diverse tissue types, representing unparalleled tissue diversity, the sample size of only 838 donors restricts generalizability.

In this study, we leverage the largest available AD GWAS summary statistics comprising over 65,000 individuals along with the largest blood and brain *cis*-eQTL summary statistics from published meta-analyses to identify novel AD-TWAS associations. Using the OTTERS approach [17], which combines diverse modeling techniques to account for variations in the sparseness or polygenicity of gene regulation, we generated candidate TWAS associations. These candidates were then rigorously filtered and validated to distill the most robust TWAS signals. Additionally, we fine-mapped causal eQTLs and performed conditional analyses to provide deeper insights into the genetic architecture underlying the identified TWAS associations.

## 2 METHODS

### 2.1 Data Resources

#### 2.1.1 Sources of Summary Statistics

We leveraged the largest available *cis*-eQTL meta-analysis summary statistics for two key AD-associated tissues:

- Cortical brain tissue (N=2,683) from the MetaBrain study [11]
- Blood (N=31,684) from the eQTLGen study [12]

We performed an inverse variance-weighted fixed-effects meta-analysis across the MetaBrain study-specific results, excluding the GTEx study, which we used for model performance evaluation. Similarly, we obtained a version of the eQTLGen meta-analysis summary statistics that excluded GTEx.

We used the largest clinically diagnosed AD meta-analysis from Kunkle et al. 2019 (Stage I: AD Cases=21,982; Controls=44,944) to perform our primary analysis so our generated results would be most reflective of an AD phenotype (**Supplementary Table 1**) [37]. However, we also performed all analyses using AD-related Dementia (ADRD) summary statistics from Bellenguez et al. 2022 as this is the largest GWAX study to date related to AD [5] and include these analyses in **Supplementary Table 2**.

We limited our analysis to European-descent populations, as all four meta-analyses were based on this population.

#### 2.1.2 Linkage Disequilibrium Reference Dataset

Because we used summary statistics, an individual-level reference dataset was needed to estimate linkage disequilibrium (LD) for model building in OTTERS and fine-mapping. We used 503 unrelated EUR samples from the 1000 Genomes high coverage release [38] (Link: https://ftp.1000genomes.ebi.ac.uk/vol1/ftp/data_collections/1000G_2504_high_coverage/working/20220422_3202_phased_SNV_INDEL_SV/) as a reference and filtered the dataset to only contain variants common to both the 1000 Genomes and GTEX (v8) (Link: https://www.gtexportal.org/home/downloads/adult-gtex/overview).

#### 2.1.3 Standardization of Summary Statistics

Both MetaBrain [11] and ADRD summary statistics [5] were available in build 38. We lifted eQTLGen [12] and AD summary statistics [37] from GRCh37/hg19 to GRCh38/hg38 using UCSC liftOver (https://hgdownload.soe.ucsc.edu/downloads.html). To ensure consistency in effect direction/coded allele, we aligned all variants to GRCh38, removing variants with a reference or alternative allele that did not match the GRCh38 reference and flipping direction of the effect as appropriate. After our alignment procedure, we also wanted to ensure that the SNPs used in expression prediction models were available in all other datasets, so we restricted our analysis to only include variants consistently available within eQTL summary statistics, AD summary statistics, 1000 Genomes, and GTEx. Using these criteria, we examined a total of 7,859,831 SNPs for the brain cortex analysis and 7,653,537 SNPs for the whole blood analysis. When using the ADRD summary statistics, we explored a total of 7,952,002 SNPs for the analysis of brain cortex analysis and 7,638,319 SNPs for the analysis of whole blood analysis.

### 2.2 OTTERS Implementation

We extended the recently published OTTERS framework [17] in several ways. In *Stage I: Training*, we used the standard five polygenic risk score (PRS) methods for summary statistic data implemented in OTTERS: p-value thresholding with LD clumping (P+T) at two thresholds (p < 0.05 & p < 0.001) [25]; elastic net regression implemented in the method *lassosum* [19]; Bayesian regression with a continuous shrinkage (CS) prior on SNP effect sizes implemented in the method PRS-CS [23]; and finally a non-parametric Bayesian multiple Dirichlet progress regression method SDPR [24] (**Figure 1**). In *Stage II: Testing*, the p-values produced from each of these modeling approaches were subsequently aggregated using the aggregated Cauchy association test (ACAT) to generate an omnibus p-value (ACAT-O) [39] (**Figure 1**). An overview of our processing and results filtering workflow is shown in **Figure 1**.

**FIGURE 1.**
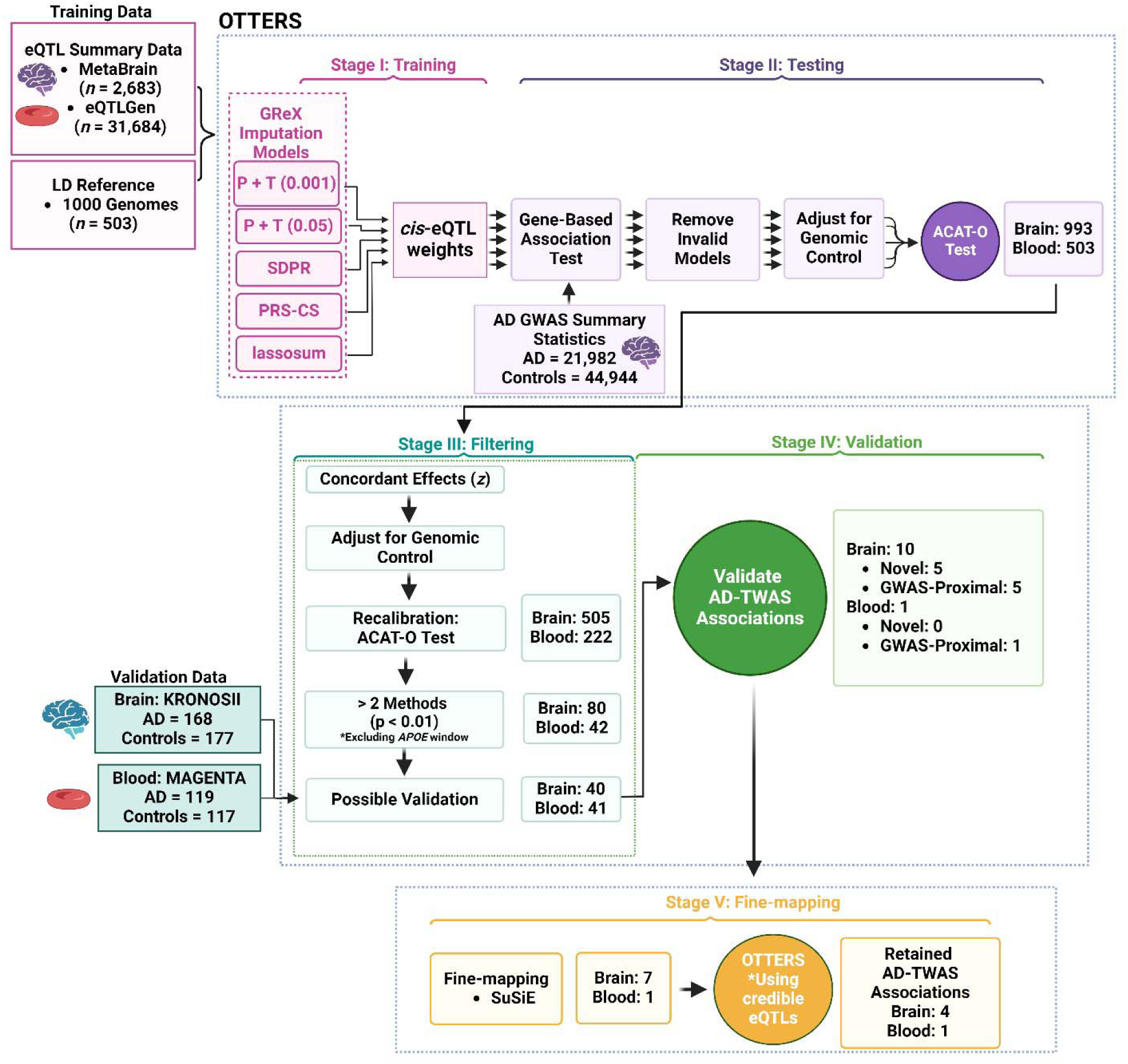
OTTERS Workflow. OTTERS uses eQTL summary data and a LD reference panel to train five imputation models (P+T 0.001, P+T 0.05, lassosum, SDPR, PRS-CS) (Stage I). These models are subsequently applied to AD GWAS summary data and the p-values of valid models are combined using ACAT-O (Stage II). We filter the results based on concordant *z* scores and validate the AD-TWAS associations using RNA-seq data (Stage III & IV). Finally, we further explore the AD-TWAS association using fine-mapping and re-running of the OTTERS workflow using only credible eQTLs to assess whether a sparse or polygenic architecture is needed for the TWAS association (Stage V). Created in BioRender.com. Abbreviations: LD, Linkage Disequilibrium; GReX, genetically-regulated gene expression; TWAS, Transcriptome-wide association study; GWAS, Genome-wide association study; AD, Alzheimer’s Disease; eQTL, expression quantitative trait loci; OTTERS, Omnibus Transcriptome Test using Expression Reference Summary data; SuSiE, Sum of Single Effects model

While the original implementation of OTTERs (https://github.com/daiqile96/OTTERS) used the *pseudovalidate* option within the *lassosum* (v.0.4.5) method to select the optimal tuning parameter between Ridge and Lasso regression, we instead used the *validate* option to generate our models with GTEX (v8) EUR normalized expression, covariate, and genetic data from Brain_Cortex and Whole_Blood (Link: https://www.gtexportal.org/home/downloads/adult-gtex) as validation sets. This modification increased model sparsity and had a higher predictive performance relative to the *pseudovalidate* option (**Supplemental Figure 1**). As we used GTEx v8 expression data to evaluate the performance of all imputation models, we prevented overfitting of the *lassosum* model by only including a randomly generated 50% split of the GTEx data (Brain: N=92/183; Blood: N=279/558) to serve as a validation testing dataset during model training and the remaining 50% served as an independent dataset for model evaluation. We also expanded the shrinkage and mixing parameters (balance between Ridge and Lasso regression) of the OTTERS *lassosum* implementation by also testing shrinkage penalties of 0.05, 0.1, 0.25, 0.33, 0.75, and 0.95 and mixing parameters of 0.05, 0.2, 0.25, 0.3, 0.4, 0.5, 0.6, 0.7, 0.75, 0.8, 0.9, and 0.95.

#### 2.2.1 Post-processing of findings from OTTERS

In the original OTTERS implementation, invalid gene models are filtered based on a *R^2^*>0.01 and after models are adjusted for genomic control. We improve upon the existing approach by instead requiring an *R*>0.1 between the predicted and actual normalized expression of the gene based on GTEx v8 data as using only the *R^2^* value masks genes models with negative correlations that are biologically nonsensical. In *Stage II: Testing*, we remove invalid models prior to adjusting for genomic control to keep them from skewing the adjustment (**Figure 1**). Overall, we used valid *z*-scores to adjust p-values for genomic control and subsequently ran the initial ACAT-O test. Multiple-testing was controlled by a Bonferroni-correction of the ACAT-O p-values identified at the end of *Stage II: Testing* (**Figure 1**). In *Stage III: Filtering*, we tested if *z*-score direction was concordant among tested methods and replaced gene models with *NA* values if there was any discordance across the methods, effectively requiring all methods to agree on the direction of effect (**Figure 1**). After we applied this concordance restriction, we applied genomic control to the concordant *z*-scores, and re-ran that ACAT-O test to yield ACAT-O_CON_ (concordance-restricted ACAT-O) p-values. By applying genomic control at this stage of the analysis, we reduced the overall ACAT inflation (Brain: λ = 1.58 > λ_CON_ = 1.17; Blood: λ = 1.27 > λ_CON_ = 1.04) (**Supplementary Figures 2-5**).

While each of the tested methods has advantages and disadvantages that lead to differing SNP selection, ultimately all methods receive the same set of input eQTL summary statistics. By enforcing concordance among the tested methods, we can enrich our results for higher confidence TWAS associations. We subsequently annotated whether a gene fell within a known AD GWAS association based on whether a 1Mb window around the gene’s transcription start and end site overlapped with a 1Mb window around *APOE* or whether a 1Mb window around the gene fell within a 1Mb window around a known AD/ADRD GWAS association. We used a combined list of lifted-over AD GWAS associations [37] and ADRD GWAS associations [5] for this annotation of the results. Finally, we restricted our associations to genes where there was at least a nominally significant p-value (p < 0.01) for a minimum of two out of five methods tested (in addition to a Bonferroni-corrected significant ACAT-O_CON_ p-value) (**Figure 1**). We implemented these criteria to minimize the potential for false-positive associations and prevent any single method from biasing our results while still allowing for the detection of multiple different model types.

#### 2.2.2 Independent Validation of OTTERS-identified Genes

In *Stage IV: Validation*, results meeting our post-processing criteria were further validated in an independent analysis of differential gene expression among AD cases and controls (**Figure 1**). We performed one-tailed T-tests (using the direction of effect predicted in our results) within a non-Hispanic white (NHW) whole-blood dataset [40] (AD Cases=119; Controls=117) and a partially independent brain RNA microarray dataset, KRONOSII [41], that was curated for high AD pathology and low secondary pathology (AD Cases=168; Controls=177). More specifically, we utilized the *RNA_eQTL_residual-corrected-data_KRONOSII_Brainome.txt* for the validation of brain cortex results (Link: https://drive.google.com/drive/folders/15WmJsBXIH3Feib9Vu_bU2VpYQ2pDc8WX).

For the whole blood validation, we used residualized normalized expression data after covariate adjustment for sex, age-at-exam, and principal components 1-12 (PC1-12) to capture population substructure estimated analysis using *flashpcaR* [42]. Because we used summary statistics, it was difficult to rule out the possibility that some samples from the KRONOSII study were included in the MetaBrain study or the AD summary statistics, however we assume based on documentation of the KRONOSII study that any overlap would be somewhat limited [41]. We used a Bonferroni-corrected p-value based on the number of genes tested to determine if a TWAS hit was validated in the actual expression data. In the microarray expression data, multiple probes mapped to a single gene in some instances, and we required at least one probe to be significantly differentially expressed between AD Cases and Controls for that the TWAS gene model to be validated.

#### 2.2.3 Fine-mapping Validated Associations

In *Stage V: Fine-mapping*, we identify the specific eQTLs driving the significant TWAS results by applying the SuSiE (Sum of Single Effects) [43,44] approach (*susieR* package v.0.12.35) to the original summary statistics while also using the same LD reference panel used in the OTTERS analysis. Credible sets of causal eQTLs were generated for eight of eleven validated TWAS associations, though seven of these eight SuSiE analyses reported convergence issues in the model fitting, which could indicate a mismatch between the eQTL summary statistics and the 1000 Genomes EUR LD matrix. For consistency however, we used the same LD matrix for all analyses. We extracted the variants falling within identified credible sets and subsequently re-ran the modified OTTERS pipeline using only the SuSiE identified SNPs to assess how the sparse fine-mapped eQTLs influence TWAS model results. We only re-ran our model for our validated TWAS associations from blood and brain. As we only performed analyses on a small set of genes, we did not adjust for genomic control and instead focused on the overall ACAT-O p-values. We annotated the credible SNPs included in the models using the Functional genomics repository (FILER) [45].

#### 2.2.4 GWAS Conditional Analysis

For our validated AD-TWAS associations that were proximal to known AD/ADRD GWAS loci, we sought to determine whether the genetically-regulated expression of the identified genes was driving the original GWAS association. To this end, we leveraged individual-level data from the Alzheimer’s Disease Genetics Consortium (ADGC), encompassing 29,681 NHW participants across 35 cohorts. We conducted a series of logistic regression models that included both the predicted genetically-regulated expression of the AD-TWAS gene and the GWAS SNP dosage, along with covariates such as sex, age, cohort, and population substructure (PC1-3). These analyses closely mirrored the approach used by Kunkle et al. 2019 [37]. When the logistic regression results warranted further investigation, we also performed linear regression models using either the GWAS SNP dosage or the genetically-regulated gene expression as the outcome variable, in order to disentangle the significant predictors. For this conditional analysis, we tested the top two most significantly associated modeling methods that produced valid gene models, with the exception of *MTCH2*, where we tested all sparse eQTL models as this was the only gene where every approach yielded a significant AD-TWAS association (**Supplementary Table 4**). Additionally, we used PLINK1.9 to perform linkage disequilibrium calculations (--r2 square) within this ADGC cohort to assess the strength of LD between AD/ADRD GWAS SNP and the causal eQTLs contained within the sparse genetically-regulated gene expression models.

## 3 RESULTS

### 3.1 OTTERS Results and Quality Filtering

For brain-cortex, 993 models out of 9,282 were significantly associated with AD (Bonferroni-corrected ACAT-O p < 5.39×10^-6^) (**Figure 1**: Stage II). We applied additional stringent filtering criteria to these prediction models, with 505 models showing concordant effect among all valid model *z*-scores (**Figure 1**: Stage III) and of those models 80 have nominal associations among at least two methods excluding five of these genes that fall within a 1Mb window of *APOE* (**Figure 1**: Stage III). For whole blood, 503 models out of 7,214 were significantly associated with AD (Bonferroni-corrected ACAT-O p < 6.93×10^-6^) (**Figure 1**: Stage II). After filtering, 222 models show concordant effect and of those models 42 have nominal associations amongst at least two individual methods excluding two of these genes that fall within a 1Mb window of *APOE* (**Figure 1**: Stage III).

### 3.2 Annotation of Significant TWAS Associations

We assumed significant TWAS associations from our OTTERS approach were driven by either 1) re-weighting and aggregating multiple sub-significant GWAS associations into a single TWAS gene association, or 2) identification of a TWAS-significant gene by either directly selecting a GWAS-associated variant in the expression prediction model or indirectly tagging a GWAS-associated variant through linkage disequilibrium. Thus, we annotated our TWAS results as either novel or GWAS-proximal, respectively, by creating 1Mb regions around variants identified as significant GWAS associations from Kunkle et al. 2019 and Bellenguez et al. 2022, and assessing whether this window overlaps with a 1Mb window around each TWAS gene [5,37]. Using this criterion, we identified 80 significant genes (excluding five genes falling within the *APOE* window) of which 50 were novel genes and 30 were GWAS association-proximal genes within brain cortex. Similarly, we identified 42 significant genes within blood (excluding two genes falling within *APOE* window) of which 18 were novel genes and 24 were GWAS association-proximal genes. Comparing results between blood and brain, there were no novel genes common between the two tissues. For genes that are proximal to known GWAS loci, we identified four genes that were identified in both whole blood and brain cortex, and which also had concordant effect directions in both tissues (*KANSL1, ARL17A, LRRC37A2*, and *C1QTNF4*) (**Supplementary Table 3**).

### 3.3 Validation in Expression Datasets

To validate TWAS associations from whole blood, we examined differences in gene expression within an independent whole-blood RNA-seq dataset of NHW, clinically-diagnosed AD cases (N=119) and age-and-sex-matched controls (N=117). Based on the direction of effect from each TWAS association, we tested if measured gene expression differed significantly between AD cases and controls using a one-tailed t-test. We were able to test 40/41 (98%) of our novel and GWAS-Proximal genes.

From this analysis, we validated one gene (*MYBPC3*) that was proximal to the known AD/ADRD GWAS locus *SPI1* (**Table 1**). Specifically, we found a positive association between *MYBPC3* (myosin-binding protein C) expression and AD risk (ACAT-O_CON_ p = 4.43×10^-8^; t-test p = 6.5×10^-4^).

**TABLE 1.**
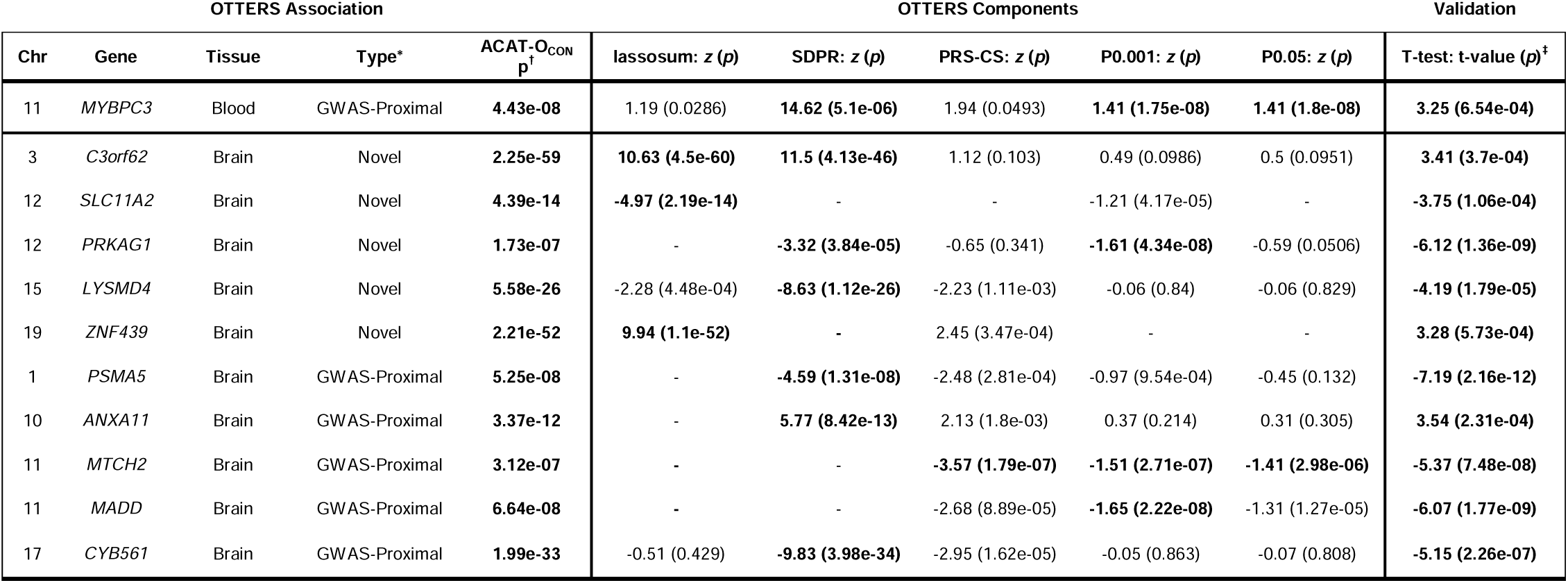
Validated TWAS associations using AD GWAS summary statistics. * Whether an association is deemed novel or GWAS-Proximal is based on whether a 1 Mb window around the transcription start and end site overlaps with a 1 Mb window around a known AD/ADRD GWAS loci. ^†^ ACAT-O_CON_ p derived from p-values generated during Stage III (Brain Cortex: Bonferroni-corrected p<5.39×10^-6^; Blood: Bonferroni-corrected p<6.93×10^-6^) (see Figure 1) ^‡^ One-tailed t-test statistics and p-values, direction of effect determined based on direction of AD-TWAS association Bold text indicates p-values significant after Bonferroni correction Abbreviations: TWAS, Transcriptome-wide association study; GWAS, Genome-wide association study; AD, Alzheimer’s Disease; ADRD, Alzheimer’s Disease and Related Dementias; OTTERS, Omnibus Transcriptome Test using Expression Reference Summary data

Brain cortex associations were validated using a brain microarray-based expression dataset of neuropathologically curated cases that were enriched for Alzheimer’s disease pathology and minimized co-occurring neurodegeneration markers such as Lewy bodies, and controls were neuropathologically confirmed as having minimal pathology loads [41]. Based on the microarray data, we were able to assess 40/80 of TWAS associations identified using the AD GWAS summary statistics. We validated ten associations. Five of 23 novel AD TWAS associations were statistically significant (Bonferroni-corrected t-test p-value<0.001; *C3orf62, LYSMD4, PRKAG1, ZNF439, SLC11A2*). In addition, we validated five AD TWAS associations (5/17) that were located proximal to a known AD/ADRD GWAS loci (Bonferroni-corrected t-test p-value < 0.001 *PSMA5, ANXA11, MTCH2, MADD, CYB561*).

### 3.4 Fine-mapping TWAS Associations

We next explored whether a sparse genetic model based on fine-mapped causal eQTLs for each *cis*-regulatory region would produce a similar strength TWAS association with improved interpretability. To test this assumption, we analyzed eQTL meta-analysis summary statistics using SuSiE and subsequently re-ran our OTTERS pipeline using only this subset of fine-mapped eQTLs (*Stage V: Fine-mapping*, **Figure 1**). Credible sets of causal eQTLs were generated for eight of eleven validated TWAS associations. Of the eight validated associations with credible sets, five genes (*MTCH2, MADD, CYB561, ZNF439,* and *MYBPC3*) had equivalent or improved AD-TWAS association statistics when the OTTERS analysis was restricted to the fine-mapped causal eQTLs within these credible sets (**Table 2**). In contrast, the remaining three genes (*C3orf62, LYSMD4,* and *ANXA11*) were no longer significant, indicating that their effects rely on a more polygenic genetic architecture than captured by the SuSiE fine-mapping (which was limited to 10 credible sets).

**TABLE 2.** Validated TWAS associations using fine-mapped causal eQTLs applied to AD GWAS summary statistics. * Whether an association is deemed novel or GWAS-Proximal is based on whether a 1 Mb window around the transcription start and end site overlaps with a 1 Mb window around a known AD/ADRD GWAS loci. ^†^ ACAT-O p derived from p-values generated during Stage V: Fine-mapping (Brain Cortex: Bonferroni-corrected p<5.39×10^-6^; Blood: Bonferroni-corrected p<6.93×10^-6^) (see Figure 1) Bold text indicates p-values significant after Bonferroni correction Abbreviations: TWAS, Transcriptome-wide association study; GWAS, Genome-wide association study; AD, Alzheimer’s Disease; ADRD, Alzheimer’s Disease and Related Dementias; eQTL, expression quantitative trait loci; OTTERS, Omnibus Transcriptome Test using Expression Reference Summary data

| OTTERS Association |  |  |  |  | OTTERS Components |  |  |  |  |
| --- | --- | --- | --- | --- | --- | --- | --- | --- | --- |
| Chr | Gene | Tissue | Type* | ACAT-O p<sup>†</sup> | lassosum: z (p) | SDPR: z (p) | PRS-CS: z (p) | P0.001: z (p) | P0.05: z (p) |
| 11 | *MYBPC3* | Blood | GWAS-Proximal | **6.48e-44** | - | - | - | **13.90 (6.48e-44)** | **13.90 (6.48e-44)** |
| 19 | *ZNF439* | Brain | Novel | **3.79e-97** | **20.92 (3.79e-97)** | - | - | - | - |
| 11 | *MTCH2* | Brain | GWAS-Proximal | **1.81e-13** | **-5.51 (3.53e-08)** | **-7.57 (3.62e-14)** | **-6.54 (6.04e-11)** | **-5.54 (3.05e-08)** | **-6.24 (4.43e-10)** |
| 11 | *MADD* | Brain | GWAS-Proximal | **1.51e-07** | -0.72 (0.474) | **-5.54 (3.01e-08)** | -3.09 (1.99e-03) | -0.16 (0.875) | -0.36 (0.716) |
| 17 | *CYB561* | Brain | GWAS-Proximal | **8.81e-20** | -2.05 (0.040) | **-9.28 (1.76e-20)** | -1.90 (0.057) | -0.66 (0.511) | -0.66 (0.509) |

#### 3.4.1 Brain: *MTCH2* & *MADD*

The *MTCH2* OTTERS association was primarily driven by the PRS-CS approach modeling 1,336 SNPs (ACAT-O_CON_ p = 3.12×10^-7^; p_PRS-CS_CON_ p = 1.79×10^-7^; *z*_PRS-CS_ = −3.57; *R^2^* = 5.7%). This model was reduced to seven credible sets with eight causal eQTLs by SuSiE fine-mapping, which produced significant AD-TWAS associations using all strategies and each method selected between one to three causal eQTLs. The most parsimonious model was generated by the lassosum approach and contained a single eQTL that falls with active enhancer/promoter regions based on chromatin accessibility and histone modification data (**Supplemental text**). This model demonstrated slightly improved model performance compared to the original (5.7% vs. 6.3% variance in gene expression explained) and maintained a strong association to AD (p_Lassosum_ = 3.53×10^-8^; z_Lassosum_ = −5.51). This variant is located approximately 235kb downstream from the established *SPI1* AD locus. In our analysis, a valid model for *SPI1* within brain was generated only using SDPR and was not significantly associated with AD (ACAT-O_CON_ p = 0.47; *z*_SDPR_ = 0.59; *R^2^* = 1.0%). We ran a conditional analysis using a series of logistic regression models within a subset of cohorts analyzed by Kunkle et al. 2019 and confirmed that there are individually significant associations between the *SPI1* locus and AD status (*z* = −3.2, p = 1.35×10^-3^) and the genetically-regulated expression of *MTCH2* and AD status (z = −4.6, p = 4.08×10^-6^). However, when both variables are included within the model only the genetically-regulated expression of *MTCH2* remains significant (z = −3.3, p = 9.09×10^-4^) whereas the *SPI1* association is no longer significant (z = −0.18, p = 0.86). These results suggest that the genetically-regulated expression of *MTCH2* appears to be a driving factor behind the *SPI1* AD GWAS locus association.

However, *MADD* is located approximately 90kb upstream from the *SPI1* GWAS association [37] and 350kb upstream from *MTCH2*. The *MADD* OTTERS association was primarily driven by the P+T(0.001) approach modeling 307 SNPs (ACAT-O_CON_ p = 6.64×10^-8^; p_P+T_0.001_CON_ = 2.22×10^-8^; *z*_P+T_0.001_ = −1.65; *R^2^*= 4.9%). This model was reduced to six credible sets with 14 potential causal eQTLs by SuSiE fine-mapping, which produced a six SNP SDPR model demonstrating slightly improved model performance to the original (4.9% vs. 6.3% variance in gene expression explained). This model also maintained a strong negative association to AD (p_SDPR_ =3.01×10^-8^; *z*_SDPR_= −5.54). Based on our earlier results with *MTCH2*, we assessed in a series of conditional models whether *MADD* contributes to the *SPI1* locus association and found that *MADD* genetically-regulated expression was not associated with AD in either the individual (z = −1.2, p=0.23) or combined models (z = −0.21, p = 0.83). Of note, we were able to impute the genetically-regulated expression of *MADD* using only five out of six SNPs within the model as the other SNP was unable to be imputed in the ADGC cohort, which may have slightly reduced the strength of the initial *MADD* association to AD. However, we confirmed using a linear model with imputed *MTCH2* expression as the outcome that the genetically-regulated expression of *MADD* (t*_MADD_* = 10.05) and the *SPI1* dosage (t*_SPI1_* = 143.35) were both significantly associated predictor variables (p < 2×10^-16^).

Moreover, the *SPI1* locus is in moderate LD with the single causal eQTL within the *MTCH2* expression model (*r*^2^ = 0.45) whereas the *MADD* SNPs are in weak LD with the *MTCH2* eQTL(*r*^2^ < 0.09) which is reflected in the strength of their effect sizes on *MTCH2* expression. Overall, these results suggest that both the *MADD* AD-TWAS association and *SPI1* locus GWAS association are driven by the *cis*-regulatory region surrounding *MTCH2*.

#### 3.4.2 Brain: *CYB561*

The *CYB561* OTTERS association was driven by the SDPR approach modeling 1,616 SNPs (ACAT-O_CON_ p = 1.99×10^-33^; p_SDPR_CON_ = 3.98×10^-34^; *z*_SDPR_ = −9.83; *R^2^* = 28.1%). Fine-mapping identified seven credible sets each containing a single causal eQTL SNP. OTTERS analysis of these SNPs identified a six SNP SDPR model with nearly identical performance (*R^2^*_SDPR_ = 24%) and produced a TWAS result of similar significance (p_SDPR_ = 1.8×10^-20^; *z*_SDPR_ = −9.3). *CYB561* is located 34kb upstream to the identified GWAS associations located near *ACE* identified in Bellenguez et al [5], and only 24kb upstream of the *ACE* association in Kunkle et al. [37]. In our analysis, all modeling approaches produced valid results for *ACE* within the brain and overall the *ACE* gene model was nominally negatively associated with AD risk (ACAT-O_CON_ p = 0.001; *z* = [-0.5, −2.6]; *R^2^* = [6%, 10%]).

As the *ACE* GWAS locus is located nearly in the middle of the promoter elements of *ACE* (∼16kb) and *CYB561* (∼24kb), we assessed in a series of logistic regression models if the genetically-regulated expression of *CYB561* is driven by its close proximity to the *ACE* GWAS locus. We found that while there was a significant association between the *ACE* locus and AD status (z = 2.9, p = 3.49×10^-3^), there was not a significant association between the genetically-regulated expression of *CYB561* and AD status (z = −0.757, p = 0.449). Moreover, we confirmed using a linear model with the dosage of the *ACE* GWAS locus as the outcome variable that the genetically-regulated expression of *CYB561* was the strongest significantly associated predictor variable (t = - 18.9, p < 2×10^-16^) despite being in weak LD with the *ACE* GWAS locus (*r*^2^ < 0.02). These results suggest that the regulatory architecture underlying this genomic region is more complex and requires further exploration in larger sample sizes to clarify the association between *CYB561* genetically-regulated expression and AD.

#### 3.4.3 Brain: *ZNF439*

The OTTERS association for *ZNF439* was driven by the lassosum approach, which modeled 1,336 SNPs. This model yielded a highly significant association to AD (ACAT-O_CON_ p = 2.21×10^-52^; p_Lassosum _CON_ = 1.1×10^-52^; *z*_Lassosum_ = 9.94; *R^2^* = 5.7%). Though causal eQTL fine-mapping, we identified five credible sets each containing a single causal eQTL and subsequent application of these SNPs to the OTTERS approach led to the generation of a four SNP lassosum model demonstrating slightly reduced model performance in terms of variance explained (*R^2^*_Lassosum_ = 5.0%) but further strengthened the association to AD (p_Lassosum_ = 3.79×10^-97^; *z*_Lassosum_ = 20.92). Functional annotation of these causal eQTLs revealed that the effect size direction of each SNP within the model corresponded to histone markers of active transcription or repressive chromatin, underscoring the additional biological complexity underlying the functional roles of these genetic variants (**Supplemental Text**).

#### 3.4.3 Blood: *MYBPC3*

The original OTTERS association to *MYBPC3* was driven most strongly by the P+T 0.001 approach modeling 1,081 SNPs (ACAT-O_CON_ p = 4.43×10^-8^; p_P+T_0.001_CON_ = 1.75×10^-8^; *z*_P+T_0.001_ = 1.41; *R^2^* = 3.6%). Fine-mapping of *MYBPC3* yielded four credible sets each containing a single causal eQTL. Restricting the OTTERS method to the credible causal eQTLs, the SDPR, lassosum, and PRS-CS modeling approaches were able to explain only a negligible proportion of gene expression, falling short of our *R^2^*>1 % filtering threshold. In contrast, the P+T methods used a two-SNP model and maintained a statistically significant positive association with AD risk (p_P+T_0.001_ = 6.48×10^-44^, *z*_P+T_0.001_ = 13.9, *R^2^* = 1%).

As *MYBPC3* is approximately 90kb upstream the *SPI1* AD GWAS locus, we also ran a conditional analysis using a series of logistic regression models to determine whether the genetically-regulated expression of *MYBPC3* within blood may be contributing to the *SPI1* locus signal. We confirmed that the *SPI1* locus was individually associated with AD status (z = −3.2, p = 1.35×10^-3^) and that there was a marginal association between the genetically-regulated expression of *MYBPC3* and AD status (z = 1.8, p = 0.07). However, when both variables were included within the model, only the *SPI1* locus association remained significant (*z* = −2.73, p = 0.006), while the *MYBPC3* genetically-regulated expression association was no longer marginally significant (*z* = −0.66, p = 0.51). Of note, we were only able to impute the genetically-regulated expression of *MYBPC3* using a single-SNP, as the other SNP could not be imputed in the ADGC cohort, which may have reduced the strength of the initial *MYBPC3* association to AD. Additionally, the single SNP within the *MYBPC3* expression model is in moderate LD with the *SPI1* GWAS variant (*r*^2^ = 0.49), which may explain the previously found marginal association to AD. Overall, these results suggest that the *MYBPC3* AD-TWAS association in blood does not contribute to the AD-GWAS signal at the *SPI1* locus.

### 3.5 Conditional Analyses in polygenic AD-TWAS Associations

We identified two genes (Brain: *PSMA5* and *ANXA11*) that were located proximal to known GWAS loci and potentially driven by polygenic eQTL architectures that were unable to be fine-mapped.

#### 3.5.1 Brain: *PSMA5*

Our identified *PSMA5* OTTERS association was driven most strongly by the SDPR approach, which modeled 2,592 SNPs (ACAT-O_CON_ p = 5.25×10^-8^; p_SDPR _CON_ = 1.31×10^-8^; *z*_SDPR_ = −4.59; *R^2^* = 3.7%), followed closely by the PRS-CS approach also modeling 2,592 SNPs (p_PRS-CS _CON_ = 2.81×10^-4^; *z*_PRS-CS_ = −2.48; *R^2^* = 5.9%). However, we were unable to fine-map any causal eQTLs within *PSMA5* using SuSiE. Given that the *SORT1* ADRD GWAS locus [5] is located approximately 66kb from the transcription start site of *PSMA5*, we assessed in a series of logistic regression models whether the *PSMA5* AD-TWAS association was driven by its proximity to the *SORT1* locus. Importantly, in our brain analyses we did not generate a valid model to predict *SORT1* expression.

We found a significant association between the *SORT1* locus and AD status (z = 6.44, p = 2.62×10^-3^) in the ADGC NHW cohort. While the SDPR model of *PSMA5* genetically-regulated expression was not significantly associated with AD (z = 1.40, p = 0.16), the PRS-CS model was significantly associated with AD (z = 2.25, p=0.024). Notably, there was minimal LD between the *SORT1* locus and the SNPs contained with the *PSMA5* model (*r*^2^ < 0.04). In the combined logistic regression model, both the *SORT1* locus and PRS-CS genetically-regulated expression of *PSMA5* retained their significant associations to AD (p_SORT1_ = 2.88×10^-3^; p_PRS-CS_PSMA5_ = 0.027), consistent with the slight LD identified between these genetic signals. Collectively, our results suggest that the identified TWAS association between *PSMA5* and AD represents a distinct genetic signal, independent of the previously reported GWAS association at the nearby *SORT1* ADRD locus.

#### 3.5.2 Brain: *ANXA11*

The *ANXA11* TWAS association was driven mostly strongly by the SDPR approach, which modeled 3,157 SNPs (ACAT-O_CON_ p = 3.37×10^-12^; p_SDPR _CON_ = 8.42×10^-13^; *z*_SDPR_ = 5.77; *R^2^*= 1.8%). Through fine-mapping, we were able to identify six credible sets containing seven causal eQTLs. However, when we re-ran the OTTERS approach restricted to these causal eQTLs, the AD-TWAS association for *ANXA11* diminished to nominal significance (p = [0.034, 0.044], despite the genetically-regulated expression explained by the model increasing from 1.8% to 4.9%. Given that *ANXA11* is located ∼343kb upstream of the *TSPAN14* ADRD GWAS locus, we assessed whether *TSPAN14* locus might be driving the *ANXA11* AD-TWAS association. Using a series of logistic regression models, we found that neither the *TSPAN14* locus nor the genetically-regulated expression of *ANXA11* was associated with AD status in this cohort. Interestingly, we did identify several SNPs within the 3,157 SNP SDPR model that were within high LD (*r*^2^ > 0.9) with the *TSPAN14* locus. Additionally, *ANXA11* was also identified as a significant ADRD-TWAS association (**Supplementary Table 2**), suggesting that the initial AD-TWAS association may be driven instead by co-occurring forms of dementia (e.g., Fronto-temporal dementia) and/or misdiagnosis. Furthermore, our findings suggest that the *TSPAN14* ADRD GWAS locus may be associated with a broader ADRD phenotype rather than a clinical AD phenotype.

## 4 DISCUSSION

In this study, we leveraged the largest available eQTL meta-analysis summary statistics from both cortical brain tissue and blood, and applied them to the largest clinically adjudicated AD case-control GWAS dataset through a summary-statistics based TWAS framework. This approach utilized five different methods to capture a range of sparse to polygenic genetic architectures underlying gene expression regulation. Using this TWAS framework, we identified and validated five novel genes within cortical brain tissue (*PRKAG1, C3orf62, LYSMD4, ZNF439, SLC11A2*) where the genetically-regulated expression of these genes was significantly associated with AD status. Additionally, we identified and validated six genes that were proximal to known AD/ADRD GWAS associations (Blood: *MYBPC3*; Brain: *MTCH2, CYB561, MADD, PSMA5, ANXA11*). Finally, we fine-mapped causal eQTLs and obtained similar or improved TWAS power for five genes (*MTCH2, MADD, CYB561, ZNF439 and MYBPC3*) when using sparse eQTL prediction models.

Our identified novel validated associations span a wide spectrum of functional understanding, with genes having limited information on function (*LYSMD4, C3orf62, ZNF439*) to genes with recognized roles in energy metabolism (*PRKAG1*) and iron homeostasis (*SLC11A2*).

*PRKAG1* (Protein Kinase AMP-activated Non-Catalytic Subunit Gamma 1) encodes a component of AMPK and is involved in sensing cellular energy. During metabolic stress, AMPK inhibits macromolecule biosynthesis and cell growth while activating energy-producing pathways. In our study, we found that low levels of *PRKAG1* within cortical brain tissue were associated with an increased AD risk (ACAT-O_CON_ p = 1.73×10^-7^). Interestingly, a 2012 study using healthy adult human cortical slices exposed to sublethal doses of amyloid-beta soluble oligomers found *PRKAG1* to be upregulated after exposure [46]. This work suggests our association may be driven by lack of proper biological compensation during high AD pathology and further work is needed to clarify the role of *PRKAG1*. While *PRKAG1* has limited associations with Alzheimer’s disease, activators of AMPK are recognized for their therapeutic potential in treating AD such as by promoting autophagy and reducing insulin resistance [47]. For instance, researchers have found that moderate aerobic exercise can counteract amyloid-beta-induced learning and memory impairment in animal models through in part AMPK restoration [48].

*SLC11A2* (Solute carrier family 11 member 2) encodes a proton-coupled divalent metal ion transporter that plays a critical role in iron homeostasis and has been previously associated with amyotrophic lateral sclerosis (ALS) and Parkinson’s disease. In our study, we found reduced expression of *SLC11A2* within the brain was associated with an increased AD risk (ACAT-O_CON_ p = 4.39×10^-14^). While there are limited studies specifically exploring the connection between *SLC11A2* and AD, one prior study used 216 AD cases and 323 controls found a nominally significant (p = 0.08) association between a variant (rs407135) within *SLC11A2* and AD [49]. Interestingly, in a prion mouse model *Slc11a2* expression within the hippocampus was found to be sustainably reduced during disease progression which is concordant with our findings [50].

Our TWAS associations identified as proximal to known AD/ADRD loci include *PSMA5* (component of 20S core proteasome complex), *ANXA11* (calcium–dependent phospholipid-binding protein), *MTCH2* (mitochondrial insertase and regulator of apoptosis and lipid homeostasis), *MADD* (adaptor protein regulating apoptosis through activation of mitogen-activated protein kinase), and *CYB561* (Transmembrane Ascorbate-Dependent Reductase).

Several previous AD TWAS studies have identified *MTCH2* as being associated with AD status, and our work further supports the notion that the genetically-regulated expression of *MTCH2* appears to drive the GWAS association at the *SPI1* locus [34,35]. Notably, we were able to retain the strength of this association using only a single causal eQTL for *MTCH2* genetically-regulated expression. This result builds upon prior work by Gockley et al. 2021 as they also identified *MTCH2* genetically-regulated expression as significantly negatively associated with AD using six neo-cortical brain tissues (N_RNA-seq = 888) [35]. Through joint-conditional probability analysis and colocalization analysis, these researchers identified that *MTCH2* likely shares a single causal variant with the *SPI1* locus [35]. However, their best performing models for *MTCH2* expression contained 260 SNPs, none of which overlapped with the SNPs identified as causal eQTLs through fine-mapping and/or retained in our OTTERS models, which highlights the importance of fine-mapping causal eQTLs using the largest available eQTL resource (**Supplementary Text**). *MADD* is also located near *SPI1* and has been found to be negatively associated AD status and our results support that this association may be driven by *MTCH2*. Critically, we cannot exclude the possibility that other nearby genes contribute to AD risk at this locus, and *SPI1* has been functionally validated with extensive modeling within myeloid cells, especially microglia, for its role in AD risk [51].

Interestingly, *ANXA11* has been found in prior work to be associated with amyotrophic lateral sclerosis (ALS) with or without frontotemporal dementia (FTD) and our work further supports the role of *ANXA11* in neurodegeneration [52]. *PSMA5* has also been associated with AD based on several *in vivo* animal models that have identified *PSMA5* downregulation is involved in APP-induced inhibition of cell proliferation, and our work further motivates additional studies to clarify how *PSMA5* expression modulation impacts AD pathogenesis [53–55]

There are several considerations to our study. First, we examined only *cis*-regulatory variants, as the OTTERS framework was designed for *cis*-eQTLs, and prior work has found that *trans*-eQTL effects tend to be relatively weaker [56]. While we used a standard 1 Mb window around the transcription start and end sites, we acknowledge that regions outside of this window may also harbor relevant eQTLs due to the complex 3D architecture of chromatin interactions.

Additionally, we conducted our analysis on cortical brain tissue, as it had the largest brain sample size available, in order to maximize statistical power for detecting tissue-specific effects, which can be challenging to disentangle using multi-tissue modeling. Prior work has suggested that approximately 8,000 samples are optimal for imputation model training in TWAS, while 56,000 samples are needed in the GWAS summary statistics to achieve maximal power [57]. Given these recommendations, we recognize that while our study represents the most well-powered AD TWAS to date through the integration of brain eQTL meta-analyses and multiple modeling approaches, larger brain sample sizes will be critical for fully elucidating the underlying genetic mechanisms of AD risk.

Although we performed fine-mapping of our TWAS associations, our results suggest only a small portion of TWAS associations are explained by fine-mapped causal eQTLs. While we propose that models not amenable to fine-mapping are likely driven by more polygenic gene expression models, it is also possible that these are explained by other effects unmeasured in our analysis, like long-range linkage disequilibrium, haplotype effects, or the tagging of structural variants or other unobserved genetic variants. Additionally, using our stringent validation criteria, we may have missed TWAS associations that are driven by gene expression changes that do not persist into later stages of the disease captured by our differential expression association analyses. However, as these results represent the best independent evaluation of our TWAS effects, we have chosen to specifically highlight genes meeting this validation criterion. Finally, our study was limited to individuals of European descent, as the eQTL meta-analysis summary statistics were generated predominantly using European-descent samples. Ongoing studies capturing greater genomic diversity will hopefully soon produce resources for similar analyses in African American and Hispanic/Latino populations.

Overall, by combining multiple large scale eQTL summary statistics with AD GWAS results, we have performed a TWAS that identified several new gene associations to AD and provided functional insights for several previously associated GWAS loci.

## Data Availability

All data produced in the present study are available upon reasonable request to the authors

## ACKNOWLEDGEMENTS

We would like all the authors who contributed to the summary statistics from the eQTL datasets (MetaBrain, eQTLGen) and AD/ADRD GWAS as these resources were invaluable to our work. We are also grateful to the authors of the OTTERS pipeline for their clear published methodology. The conditional analyses utilized data from the Alzheimer’s Disease Genetics Consortium (ADGC) (https://www.adgenetics.org/). The researchers associated with the ADGC (**Collaborators Appendix**) contributed to the planning and execution of the ADGC, and/or provided data, but were not involved in the analysis or writing of this article. The full acknowledgement statement for the Alzheimer’s Disease Genetics Consortium can be found here: https://www.adgenetics.org/content/acknowledgements. We are grateful to the contributors who gathered the samples utilized in this study, as well as the patients and their families, whose assistance and participation enabled this work to be carried out.

## CONSENT STATEMENT

All study procedures were approved by the institutional review boards at each corresponding study center and written informed consent was obtained either from the study participant or legal guardian.

## Collaborators Appendix

The collaborators associated with the Alzheimer’s Disease Genetics Consortium (ADGC) include the following individuals: Erin Abner, PhD; Perrie M. Adams, PhD; Alyssa Aguirre, LCSW; Marilyn S. Albert, PhD; Roger L. Albin, MD; Mariet Allen, PhD; Lisa Alvarez, Howard Andrews, PhD; Liana G. Apostolova, MD; Steven E. Arnold, MD; Sanjay Asthana, MD; Craig S. Atwood, PhD; Gayle Ayres, DO; Robert C. Barber, PhD; Lisa L. Barnes, PhD; Sandra Barral, PhD; Jackie Bartlett, PhD; Thomas G. Beach, MD PhD; James T. Becker, PhD; Gary W. Beecham, PhD; Penelope Benchek, PhD; David A. Bennett, MD; John Bertelson, MD; Sarah A. Biber, PhD; Thomas D. Bird, MD; Deborah Blacker, MD; Bradley F. Boeve, MD; James D. Bowen, MD; Adam Boxer, MD, PhD; James B. Brewer, MD; James R. Burke, MD PhD; Jeffrey M. Burns, MD MS; William S. Bush, PhD; Joseph D. Buxbaum, PhD; Goldie Byrd, PhD; Laura B. Cantwell, MPH; Chuanhai Cao, PhD; Cynthia M. Carlsson, MD; Minerva M. Carrasquillo, PhD; Kwun C. Chan, PhD; Scott Chasse, PhD; Yen-Chi Chen, PhD; Marie-Francoise Chesselet, PhD; Nathaniel A. Chin, MD; Helena C. Chui, MD; Jaeyoon Chung, PhD; Suzanne Craft, PhD; Paul K. Crane, MD MPH; Marissa Cranney, BS; Carlos Cruchaga, PhD; Michael L. Cuccaro, PhD; Jessica Culhane, PhD; C. Munro Cullum, PhD; Eveleen Darby, MA MS; Barbara Davis, MA; Philip L. De Jager, MD PhD; Charles DeCarli, MD; John C. DeToledo, MD; Dennis W. Dickson, MD; Nic Dobbins, PhD; Ranjan Duara, MD; Nilufer Ertekin-Taner, MD PhD; Denis A. Evans, MD; Kelley M. Faber, MS; Thomas J. Fairchild, PhD; Daniele Fallin, PhD; Kenneth B. Fallon, MD; David W. Fardo, PhD; Martin R. Farlow, MD; John Farrell, PhD; Lindsay A. Farrer, PhD; Victoria Fernandez-Hernandez, Tatiana M. Foroud, PhD; Matthew P. Frosch, MD PhD; Douglas R. Galasko, MD; Adriana Gamboa, BS; Kathryn M. Gauthreaux, PhD; Tamar Gefen, PhD; Daniel H. Geschwind, MD PhD; Bernardino Ghetti, MD; John R. Gilbert, PhD; Alison M. Goate, D.Phil; Thomas Grabowski, MD; Neill R. Graff-Radford, MD; Anthony R. Griswold, PhD; Jonathan L. Haines, PhD; Hakon Hakonarson, MD PhD; Kathleen Hall, PhD; James R. Hall, PhD; Ronald L. Hamilton, MD; Kara L. Hamilton-Nelson, MPH; Xudong Han, PhD; Oscar Harari, PhD; John Hardy, PhD; Lindy E. Harrell, MD PhD; Elizabeth Head, PhD; Victor Henderson, MD MS; Michelle Hernandez, BS; Lawrence S. Honig, MD PhD; Ryan M. Huebinger, PhD; Matthew J. Huentelman, PhD; Christine M. Hulette, MD; Bradley T. Hyman, MD PhD; Linda Hynan, PhD; Laura Ibanez, BS; Gail P. Jarvik, MD PhD; Suman Jayadev, MD; Lee-Way Jin, MD PhD; Kimberly Johnson, MSW PhD; Leigh Johnson, PhD; Bruce Jones, PhD; Gyungah Jun, PhD; M. Ilyas Kamboh, PhD; Moon Il Kang, PhD; Anna Karydas, BA; Mindy J. Katz, MPH; John S.K. Kauwe, PhD; Jeffrey A. Kaye, MD; C. Dirk Keene, MD PhD; Benjamin Keller, PhD; Aisha Khaleeq, MD; Ronald Kim, MD; Janice Knebl, DO; Neil W. Kowall, MD; Joel H. Kramer, PsyD; Walter A. Kukull, PhD; Brian W. Kunkle, PHD MPH; Amanda P. Kuzma, MS; Frank M. LaFerla, PhD; James J. Lah, MD PhD; Eric B. Larson, MD MPH; Melissa Lerch PhD; Alan J. Lerner MD; Yuk Ye Leung, PhD; James B. Leverenz, MD; Allan I. Levey, MD PhD; Andrew P. Lieberman, MD PhD; Richard B. Lipton, MD; Oscar L. Lopez, MD; Kathryn L. Lunetta, PhD; Constantine G. Lyketsos, MD MHS; Douglas Mains, DrPH; Jennifer Manly, PhD; Logue Mark, PhD; David Marquez,PhD; Daniel C. Marson, JD PhD; Eden R. Martin, PhD; Eliezer Masliah, MD; Paul Massman, PhD; Arjun V. Masurkar, MD PhD; Richard Mayeux, MD; Wayne C. McCormick, MD MPH; Susan M. McCurry, PhD; Stefan McDonough, PhD; Ann C. McKee, MD; Marsel Mesulam, MD; Jesse Mez, PhD; Bruce L. Miller, MD; Carol A. Miller, MD; Charles Mock, PhD; Abhay Moghekar, MD; Thomas J. Montine, MD PhD; Edwin Monuki, Sean D. Mooney, PhD; John C. Morris, MD; Shubhabrata Mukherjee, PhD; Amanda J. Myers, PhD; Adam C. Naj, PhD; Trung Nguyen, PhD; James Noble, PhD; Kelley Nudelman, PhD; Sid E. O’Bryant, PhD; Kyle Ormsby, PhD; Marcia Ory, PhD MPH; Raymond Palmer, PhD; Joseph E. Parisi, MD; Henry L. Paulson, MD PhD; Valory Pavlik, PhD; David Paydarfar, MD; Victoria Perez, BS; Margaret A. Pericak-Vance, PhD; Ronald C. Petersen, MD PhD; Marsha Polk, BS; Liming Qu, MS; Mary Quiceno, MD; Joseph F. Quinn, MD; Ashok Raj, MD; Farid Rajabli, PhD; Vijay Ramanan, PhD; Eric M. Reiman, MD; Joan S. Reisch, PhD; Christiane Reitz, MD PhD; John M. Ringman, MD; Erik D. Roberson, MD PhD; Monica Rodriguear, MA; Ekaterina Rogaeva, PhD; Howard J. Rosen, MD; Roger N. Rosenberg, MD; Donald R. Royall, MD; Mary Sano, PhD; Andrew J. Saykin, PsyD; Gerard D. Schellenberg, PhD; Julie A. Schneider, MD; Lon S. Schneider, MD; William W. Seeley, MD; Richard M. Sherva, PhD; Dean K. Shibata, PhD; Scott Small, MD; Amanda G. Smith, MD; Janet Smith, BS; Yeunjoo Song, PhD; Salvatore Spina, MD; Peter St George-Hyslop, MD FRCP; Robert A. Stern, PhD; Alan Stevens, PhD; Stephen Strittmatter, MD PhD; David Sultzer, BS; Russell H. Swerdlow, MD; Andrew Teich, PhD; Jeffrey Tilson, PhD; Giuseppe Tosto, MD; John Q. Trojanowski, MD PhD; Juan C. Troncoso, MD; Debby W. Tsuang, MD; Otto Valladares, MS; Vivianna M. Van Deerlin, MD PhD; Christopher Van Dyck, MD; Linda J. Van Eldik, PhD; Jeffery M. Vance, MD PhD; Badri N. Vardarajan, MS; Robert Vassar, PhD; Harry V. Vinters, MD; Li-San Wang, PhD; Sandra Weintraub, PhD; Kathleen A. Welsh-Bohmer, PhD; Nick Wheeler, PhD; Ellen Wijsman, PhD; Kirk C. Wilhelmsen, MD PhD; Benjamin Williams, MD; Jennifer Williamson, MS; Henrick Wilms, MD; Thomas S. Wingo, MD; Thomas Wisniewski, MD; Randall L. Woltjer, MD PhD; Martin Woon, PhD; Steven G. Younkin, MD PhD; Lei Yu, PhD; Yi Zhao, MS; Xiongwei Zhou, PhD; Congcong Zhu, PhD.

