## Supplemental Text for "Brain and Blood Transcriptome-Wide Association Studies Identify Five Novel Genes Associated with Alzheimer’s Disease"

*1 Causal eQTL Fine-mapping Functional Annotations*

*1.1* Brain: *MTCH2*

We retained the strength of our initially identified AD-TWAS association with *MTCH2* genetically-regulated expression using the causal eQTLs identified by SuSiE [1] fine-mapping and later applied to the OTTERS pipeline. While all OTTERS methods produced models that were significantly associated with AD, the most parsimonious model was generated by the lassosum method which retained only a single causal eQTL ($p_{Lassosum}$ = 3.53×$10^{-8}$; $z_{Lassosum}$ = -5.51). This causal eQTL (chr11_47594246_G_C_b38; Effect Size = 0.30) was positively associated with *MTCH2* expression and falls with several epigenomic marks associated with active enhancers and promoters including H3K4me1, H3K4me3, H3K9ac, H3K27ac, and regions of open-chromatin (DNase-seq, ATAC-seq, ChIA-PET CTCF peaks) based on FILER annotations.

Previously, Gockley et al. 2021 found *MTCH2* genetically-regulated gene expression was negatively associated with AD risk (Model: bslmm; $z_{TWAS}$ = -5.73; $p_{TWAS}$ = 1.02×$10^{-8}$) [2]. In this study, researchers used six neo-cortical brain tissues (N_RNA-seq = 888) and five different genetically-regulated expression modeling approaches (blup, bslmm, lasso, top1, and enet) for Stage I of the AD-TWAS. Notably, the training sample used in this prior study is approximately 34% of the size of the MetaBrain Brain_Cortex dataset (N = 2,683) [2,3]. We further investigated whether any of the identified causal eQTLs either identified by SuSiE and/or later selected by the OTTERS modeling methods using the fine-mapped causal eQTLs overlapped with the reported variants included in the models produced by Gockley et al. 2021 [2]. We lifted down the credible causal eQTLs identified initially using SuSiE fine-mapping and the set of variants selected by the OTTERS modeling approaches using the UCSC browser (https://genome.ucsc.edu/cgi-bin/hgLiftOver) with the original assembly as GRCh38/hg38 and the new assembly GRCh37/hg19. We downloaded the available models (GRCh37) from this prior work using the following link (https://adknowledgeportal.synapse.org/Explore/Studies/DetailsPage?Study=syn22313785). For *MTCH2*, none of the identified causal eQTLs initially identified by SuSiE and/or later used by the OTTERS approach were found within any of the five models produced by Gockley et al. 2021. These results illustrate the importance of fine-mapping causal eQTL variants to understand the key variants driving TWAS associations.

*1.2* Brain: *CYB561*

The *CYB561* OTTERS association was driven originally by the SDPR approach modeling 1,616 SNPs (ACAT-$O_{CON}$ $p = 1.99\times10^{-33}$; $p_{SDPR\_CON} = 3.98\times10^{-34}$; $z_{SDPR} = -9.83$; $R^2_{SDPR} = 28.1\%$). Fine-mapping identified seven credible sets each containing a single causal eQTL SNP. OTTERS analysis of these SNPs identified a six SNP SDPR model with nearly identical performance ($R^2_{SDPR} = 24\%$) and produced a TWAS result of similar significance ($p_{SDPR} = 1.8\times10^{-20}$; $z_{SDPR} = -9.3$) We further explored the functional annotations associated with the six SNPs within the SDPR model. There were two SNPs (chr17_62990456_A_T_b38 & chr17_63325078_G_A_b38) that were positively associated with *CYB561* expression and these SNPs shared H3K4me1 marks indicating active or poised enhancers. In comparison, the remaining four SNPs were negatively associated with the expression of *CYB561* and while they all shared

H3K36me3 histone marks, which are general indicators of active transcription and prevention of spurious transcription initiation, they also all shared markers of heterochromatin with histone marks of either H3K9me3 (chr17_63111035_G_A_b38, chr17_63183402_G_A_b38, and chr17_63222967_C_T_b38) or H3K27me3 (chr17_63323907_G_A_b38). Overall, these results highlight the biological complexity that may be underlying why these variants are associated with changes in *CYB561* expression.

*1.3* Brain: *ZNF439*

The OTTERS association for *ZNF439* was driven by the lassosum approach, which originally modeled 1,336 SNPs. This model yielded a highly significant association to AD (ACAT-$O_{CON}$ p = $2.21\times10^{-52}$; $p_{Lassosum\_CON} = 1.1\times10^{-52}$; $z_{Lassosum} = 9.94$; $R^2_{Lassosum}$ = 5.7%). Through causal eQTL fine-mapping, we identified five credible sets each containing a single causal eQTL and subsequent application of these SNPs to the OTTERS approach led to the generation of a four SNP lassosum model demonstrating slightly reduced model performance in terms of variance explained ($R^2_{Lassosum}$ = 5.0%) but further strengthened the association to AD ($p_{Lassosum} = 3.79\times10^{-97}$; $z_{Lassosum} = 20.92$). We further explored the functional annotations associated with the four SNPs within the lassosum model. There were two SNPs (chr19_11890134_C_T_b38 & chr19_11896203_G_C_b38) that were positively associated with *ZNF439* expression and these SNPs shared H3K36me3 marks indicative of active transcription. In addition, chr19_11890134_C_T_b38 was also enriched for several other markers of active enhancers and promoters such as H3K27ac, H3K4me1, H3K4me3, and H3K9ac. In comparison, the remaining two SNPs were negatively associated with the expression of

*ZNF439* and all shared H3K9me3 markers which are indicative of transcriptionally repressive heterochromatin regions. Of note, one of the positively associated SNPs (chr19_11896203_G_C_b38) also had annotations for H3K9me3 indicating the epigenomic regulation of this SNP is complex.

*1.4* Blood: *MYBPC3*

Fine-mapping of *MYBPC3* yielded four credible sets each containing a single causal eQTL. Restricting the OTTERS method to the credible causal eQTLs, the SDPR, lassosum, and PRS-CS modeling approaches were able to explain only a negligible proportion of gene expression, falling short of our $R^2$>1 % filtering threshold. In contrast, the P+T methods used a two-SNP model and maintained a statistically significant positive association with AD risk ($p_{P+T\_0.001}=6.48\times10^{-44}$, $z_{P+T\_0.001}=13.9$, $R^2_{P+T\_0.001}=1\%$). In terms of the functional relevance of these SNPs, chr11_47403380_G_A_b38 was positively associated with the expression of *MYBPC3* and this location corresponds with several epigenetic markers of active enhancers/promoters such as H3K27ac, H3K4me1, H3K9ac. The second SNP chr11_47433036_G_A_b38 selected by the P+T approaches was negatively associated with the expression of *MYBPC3* and, while this location is associated with heterochromatin marks (i.e., H3K9me3), we also found some markers of active transcription (H3K4me3, H3K36me3), which suggests location is not sufficient alone to understand the full functional impact of this SNPs. Overall, this result illustrated how fine-mapping the causal eQTLs within a TWAS association could lead sparse genetic models that were sufficiently powered to generate a similar strength TWAS associations that were otherwise lost by the original method.

### 2 *Elucidating the role of AD-TWAS associations*

#### *2.1* Brain: *CYB561*

*CYB561* (Cytochrome B561) is a transmembrane monodehydroascorabate reductase that serves a vital role in ascorbate-dehydroascorbate recycling within catecholamine secretory vesicles in order to produce norepinephrine (NE) from dopamine [6]. In our study, we found a negative association between the genetically regulated expression of *CYB561* and AD risk (ACAT-$O_{CON}$ $p = 1.99\times10^{-33}$) and validated that significantly lower levels of *CYB561* were found in AD brains relative to controls (t-test $p=2.26\times10^{-7}$). In terms of the functional roles of *CYB561*, individuals with homozygous pathogenic mutations within *CYB561* have been found to present with life-threatening orthostatic hypotension, recurrent hypoglycemia, and low levels of norepinephrine [6]. Interestingly, knockout *CYB561* mice displayed significantly reduced whole brain levels of norepinephrine [6]. In addition, another study found using the Drosophila mutant *nemy*, which is the *CYB561* homologue, that reduced levels of *nemy* lead to learning and memory deficits [7]. Previous studies have supported the role of NE in AD, as researchers have found that the locus coeruleus (LC) which produces NE is one of the first brain regions that degenerates during early stages of AD and that this loss corresponds with memory impairment [8]. Moreover, several *in vitro* studies have found decreased amyloid-beta toxicity and increased cell viability in the presence of dopamine and NE which further supports the therapeutic potential of NE modulation in AD [9]. Overall, our results provide an additional line of evidence to support the mechanistic role of NE in AD risk and motivate further studies to assess the role of *CYB561* expression in AD pathogenesis.

*2.2* Brain: *C3orf62*

*C3orf62* (Chromosome 3 Open Reading Frame 62) is a protein-coding gene associated with spermatogenesis and male fertility. In our study, we found a positive association between the genetically regulated expression of *C3orf62* and AD risk (ACAT-$O_{CON}$ p=$2.25\times10^{-59}$; $z_{LASSOSUM}$ = 10.63; $R^2_{LASSOSUM}$ = 2.3%) and validated that significantly higher expression levels of *C3orf62* were found in AD brains relative to controls (Mean Difference [Case vs. Controls]=0.024; t-test p=$3.7\times10^{-4}$). While little is known about the role of *C3orf62*, one study exploring the spatiotemporal brain transcriptome through various states of human development found *C3orf62* to exhibit female-sex biased differential gene expression in the dorsolateral prefrontal cortex and medial prefrontal cortex during prenatal development [10]. In another study, two TWAS approaches PrediXcan and MetaXcan identified significant associations between the genetically-regulated expression of *C3orf62* and the chronic inflammatory condition Crohn's Disease (CD) [11]. Finally, when we searched the GWAS catalog, several associations between variants within or near *C3orf62* are associated with depression. Overall, the limited information available about *C3orf62* requires further *in vitro* and *in vivo* exploration for how its expression may be related to AD.

*2.3* Brain: *LYSMD4*

*LYSMD4* (LysM Domain Containing 4) is predicted to encode for an integral membrane component. In our study, we found a negative association between the genetically regulated expression of *LYSMD4* and AD risk (ACAT-$O_{CON}$ p = $5.58\times10^{-26}$; $z_{SDPR}$ = -8.63; $R^2_{SDPR}$ = 17.5%) and validated that significantly lower expression levels of *LYSMD4* were found in AD brains relative to controls (Mean Difference [Case vs. Controls] = -

0.02; t-test $p=1.79\times10^{-5}$). There is little known about the role of *LYSMD4* in the brain. One study by Lorenzi et al. 2018 explored the functional genetic mechanisms of brain atrophy in AD and identified a SNP (rs1808723) within *LYSMD4* that was negatively associated within tibial nerve tissue to brain atrophy ($p=2.9\times10^{-24}$; Effect size = -0.66; SE=0.057) [12]. Another study performed an epigenome-wide association of stroke outcome and found a nominally significant association ($p=9.75\times10^{-6}$) between a CpG methylation site within 200 bp of the transcription start site of *LYSMD4* and stroke outcome during the discovery phase of the analysis (N=643), but this finding did not replicate (N=62) [13]. Overall, the role of *LYSMD4* in the brain remains unclear. Therefore, further studies are needed to elucidate the function of *LYSMD4* and its association with AD.

### SUPPLEMENTAL TEXT REFERENCES

[6] van den Berg MP, Almomani R, Biaggioni I, van Faassen M, van der Harst P, Silljé HHW, et al. Mutations in CYB561 Causing a Novel Orthostatic Hypotension

Syndrome. Circ Res 2018;122:846–54. https://doi.org/10.1161/CIRCRESAHA.117.311949.
