## Supplemental Figures for "Brain and Blood Transcriptome-Wide Association Studies Identify Five Novel Genes Associated with Alzheimer’s Disease"

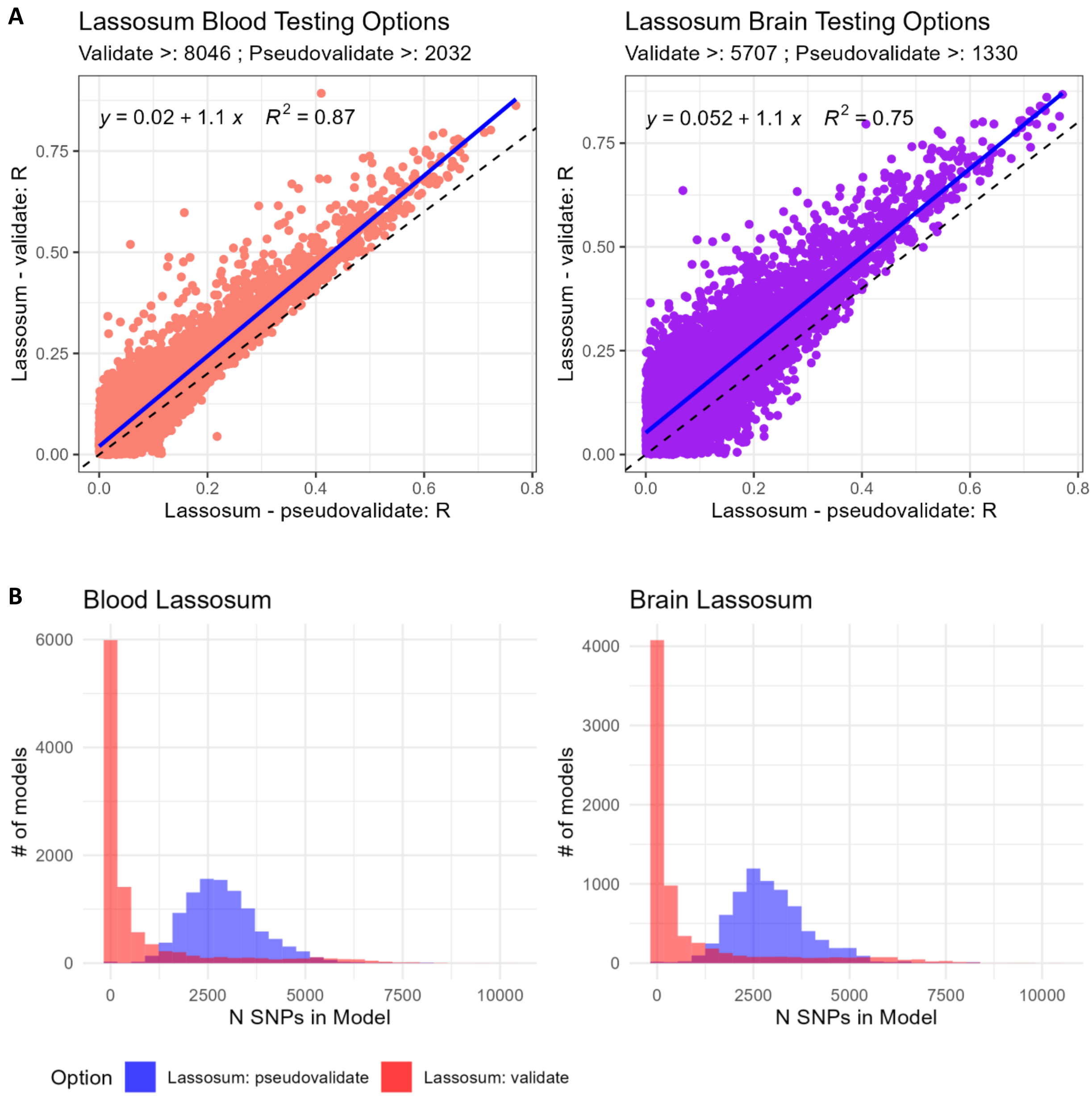


**Supplementary Figure 1**. Improved model sparsity and performance using *validate* within lassosum. We compare within blood and cortical brain tissue how the correlation between predicted and observed expression in GTEx v8 testing datasets differs between the approaches and observe increased performance using *validate* (A). We also show greatly increased model sparsity using GTEx v8 testing data as a validation dataset for hyperparameter tuning with using validate rather than *pseudovalidate* (B).

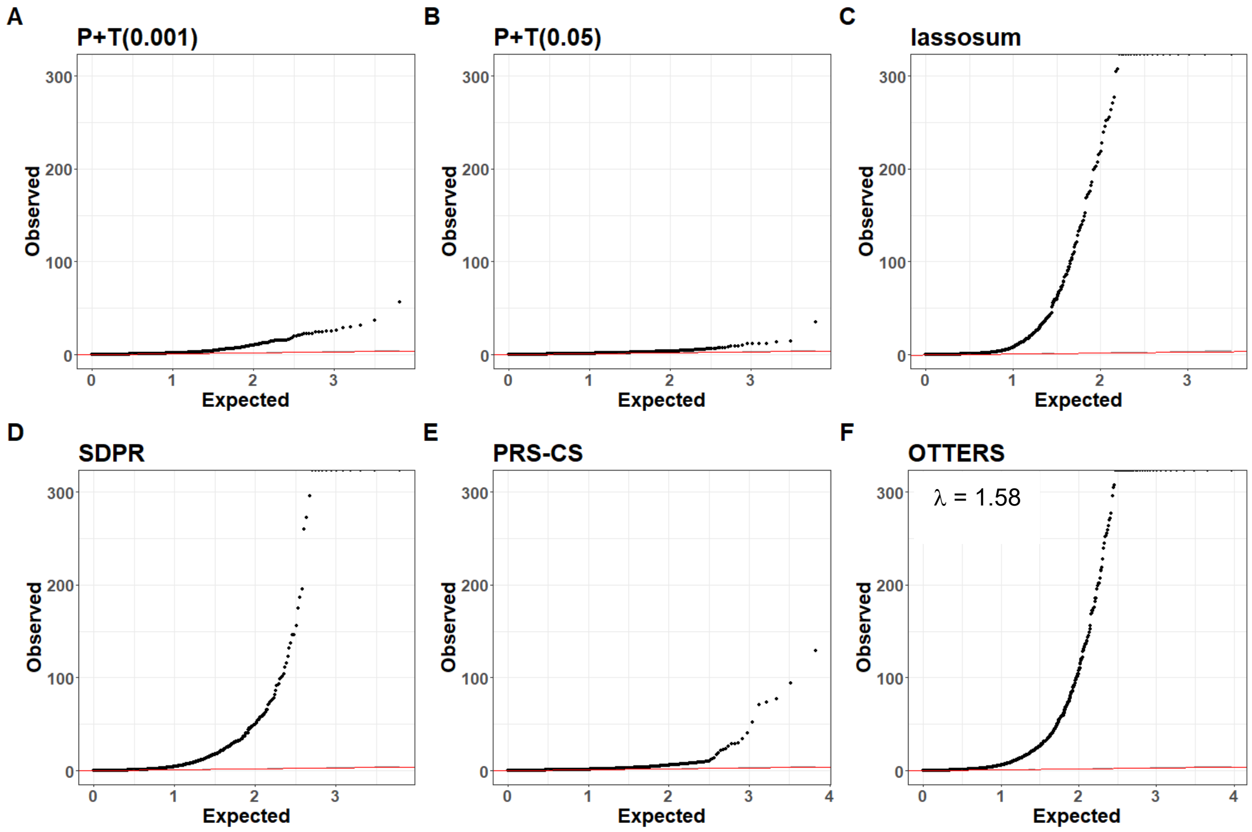


**Supplemental Figure 2.** Quantile-Quantile (QQ) plots of Brain-Cortex TWAS p-values applied to Alzheimer's Disease (AD) GWAS data of 6,424 genes by P+T(0.001) (A), 6,238 genes by P+T(0.05) (B), 3,143 genes by lassosum (C), 6,114 genes by SDPR (D), 6,634 genes by PRS-CS (E), and 9,282 genes by OTTERS (ACAT) (F). P-values are genomic-control corrected p-values generated from TWAS Z-score tests (two-sided). Only genes with valid models (Testing R > 0.1) are included within this plot.

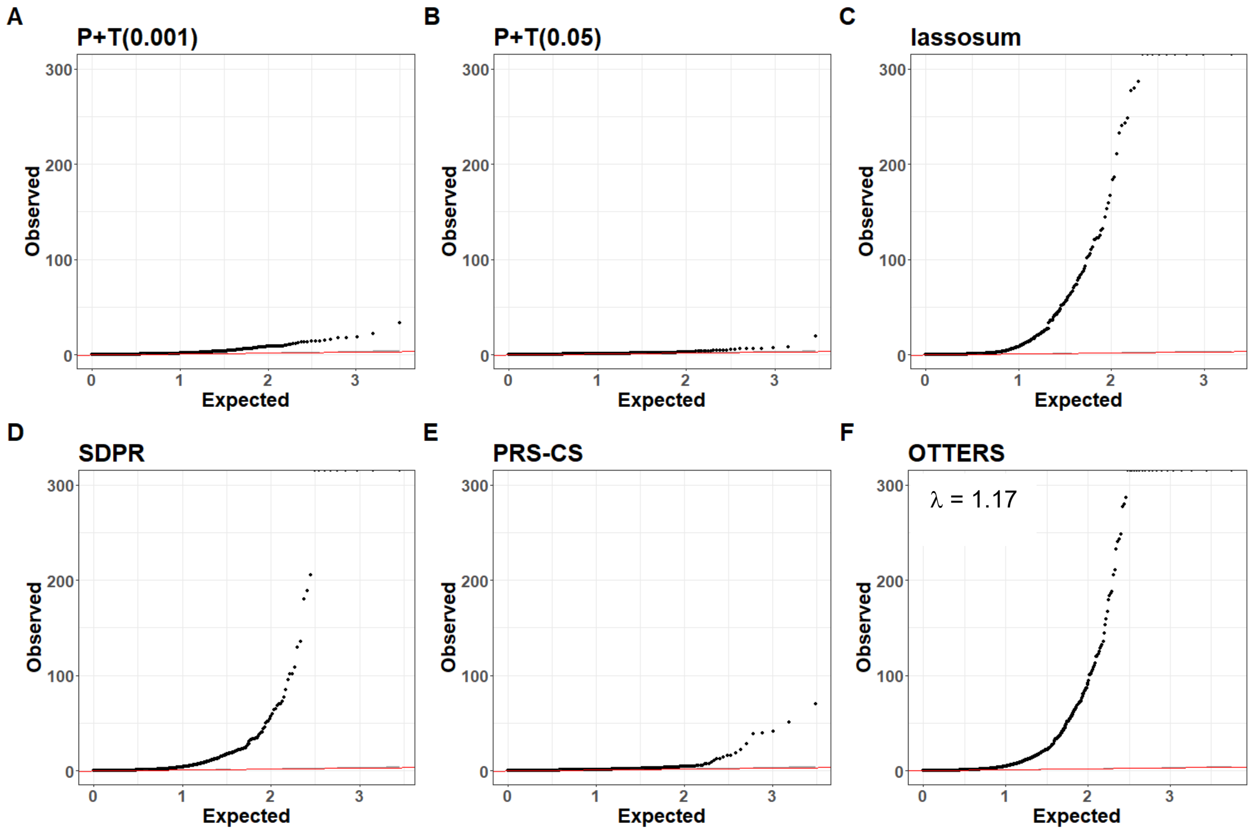


**Supplemental Figure 3.** Quantile-Quantile (QQ) plots of Brain-Cortex TWAS p-values applied to Alzheimer's Disease (AD) GWAS data of 3,115 genes by P+T(0.001) (A), 2,856 genes by P+T(0.05) (B), 1,953 genes by lassosum (C), 2,812 genes by SDPR (D), 3,060 genes by PRS-CS (E), and 5,535 genes by OTTERS (ACAT) (F). P-values are genomic-control corrected p-values generated from TWAS Z-score tests (two-sided) using only gene models with concordant effects across all modeling approaches. Only genes with valid models (Testing R > 0.1) are included within this plot.

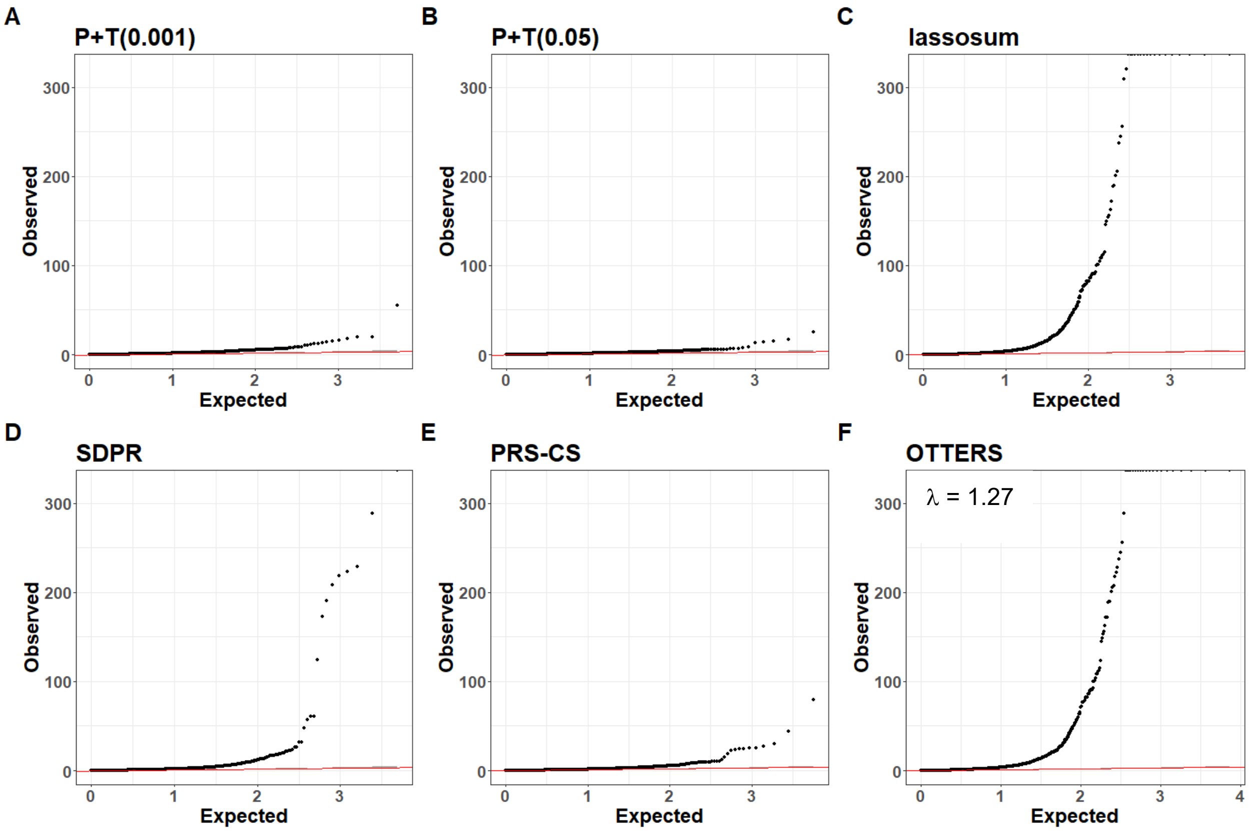


**Supplemental Figure 4.** Quantile-Quantile (QQ) plots of Blood TWAS p-values applied to Alzheimer's Disease (AD) GWAS data of 5,144 genes by P+T(0.001) (A), 4,979 genes by P+T(0.05) (B), 5,243 genes by lassosum (C), 4,830 genes by SDPR (D), 5,619 genes by PRS-CS (E), and 7,214 genes by OTTERS (ACAT) (F). P-values are genomic-control corrected p-values generated from TWAS Z-score tests (two-sided). Only genes with valid models (Testing R > 0.1) are included within this plot.

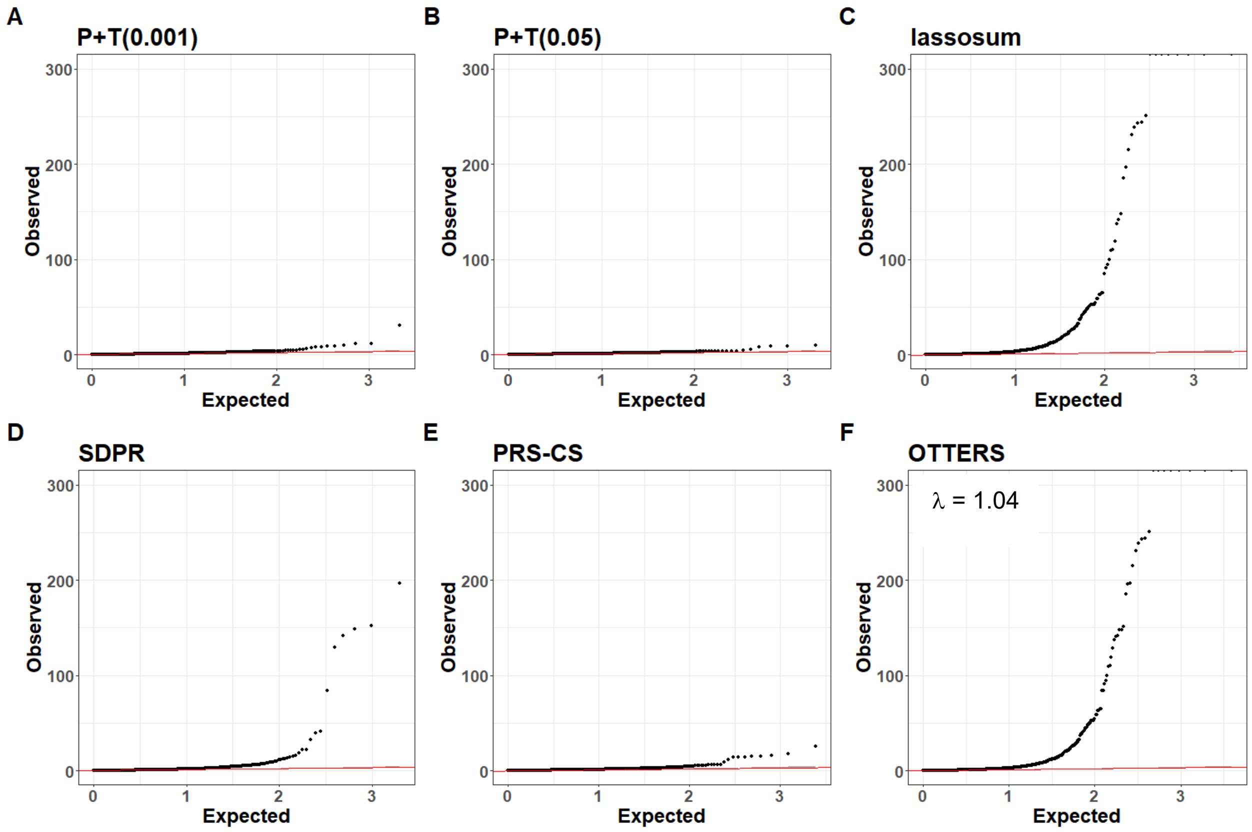


**Supplemental Figure 5.** Quantile-Quantile (QQ) plots of Blood TWAS p-values applied to Alzheimer's Disease (AD) GWAS data of 2,112 genes by P+T(0.001) (A), 2,008 genes by P+T(0.05) (B), 2,585 genes by lassosum (C), 1,972 genes by SDPR (D), 2,430 genes by PRS-CS (E), and 3,898 genes by OTTERS (ACAT) (F). P-values are genomic-control corrected p-values generated from TWAS Z-score tests (two-sided) using only gene models with concordant effects across all modeling approaches. Only genes with valid models (Testing R > 0.1) are included within this plot.

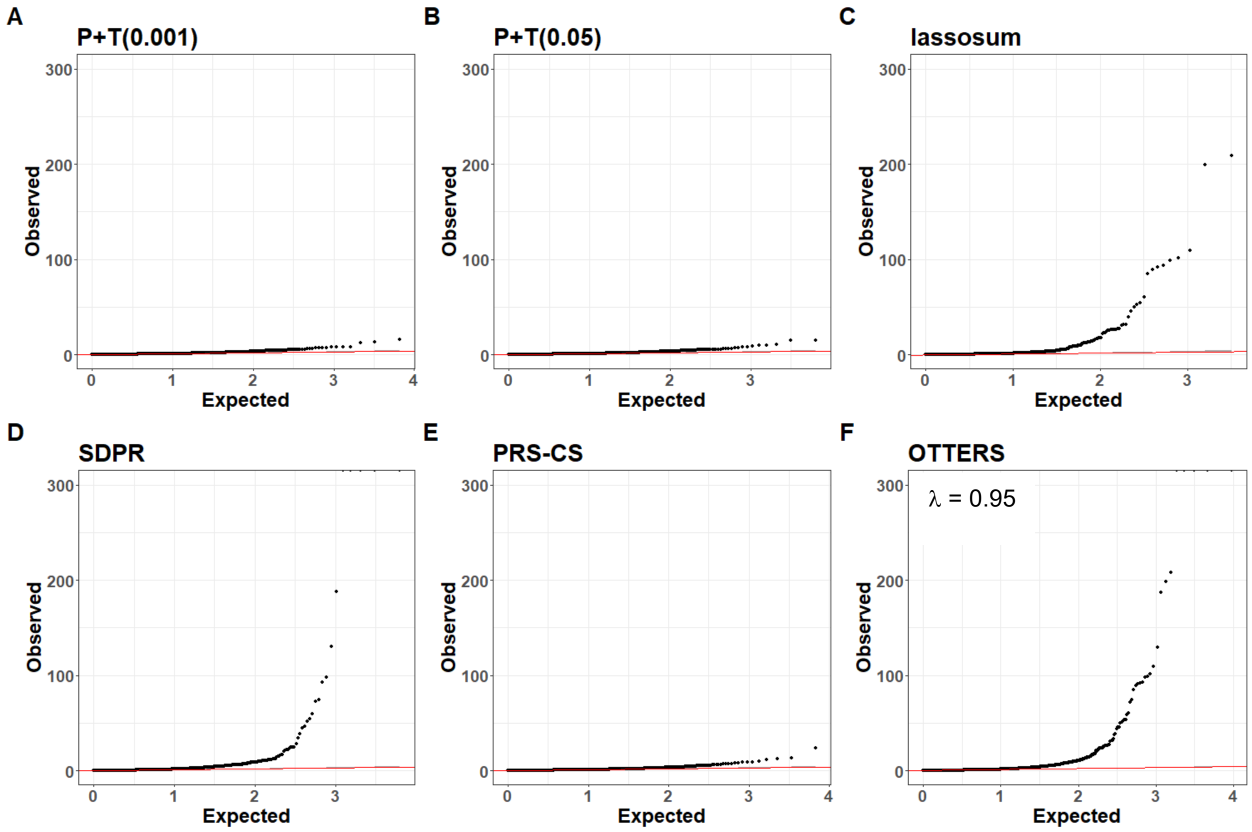


**Supplemental Figure 6.** Quantile-Quantile (QQ) plots of Brain-Cortex TWAS p-values applied to Alzheimer's Disease and Related Dementias (ADRD) GWAS data of 6,547 genes by P+T(0.001) (A), 6,342 genes by P+T(0.05) (B), 3,180 genes by lassosum (C), 6,186 genes by SDPR (D), 6,754 genes by PRS-CS (E), and 9,394 genes by OTTERS (F). P-values are genomic-control corrected p-values generated from TWAS Z-score tests (two-sided). Only genes with valid models (Testing R > 0.1) are included within this plot.

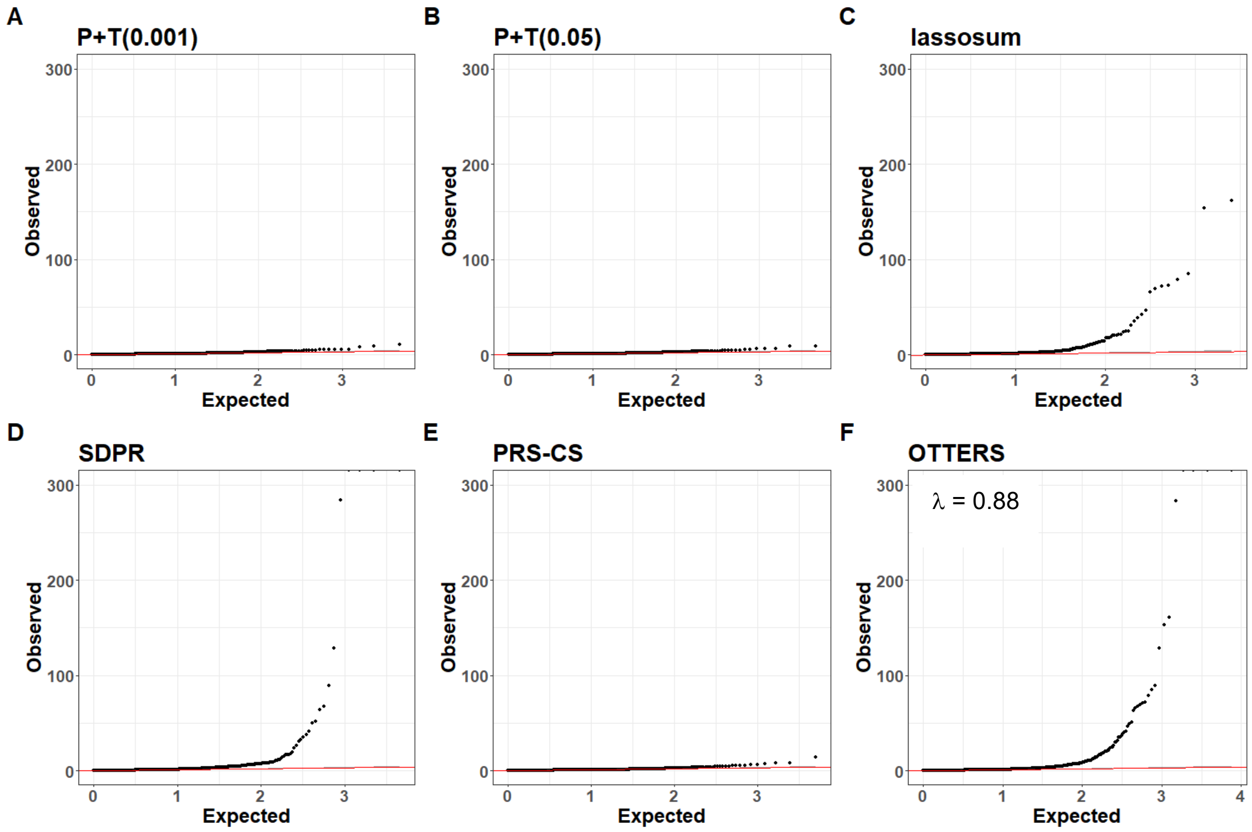


**Supplemental Figure 7.** Quantile-Quantile (QQ) plots of Brain-Cortex TWAS p-values applied to Alzheimer's Disease and Related Dementias (ADRD) GWAS data of 6,547 genes by P+T(0.001) (A), 6,342 genes by P+T(0.05) (B), 3,180 genes by lassosum (C), 6,186 genes by SDPR (D), 6,754 genes by PRS-CS (E), and 9,394 genes by OTTERS (F). P-values are genomic-control corrected p-values generated from TWAS Z-score tests (two-sided) using only gene models with concordant effects across all modeling approaches. Only genes with valid models (Testing R > 0.1) are included within this plot.

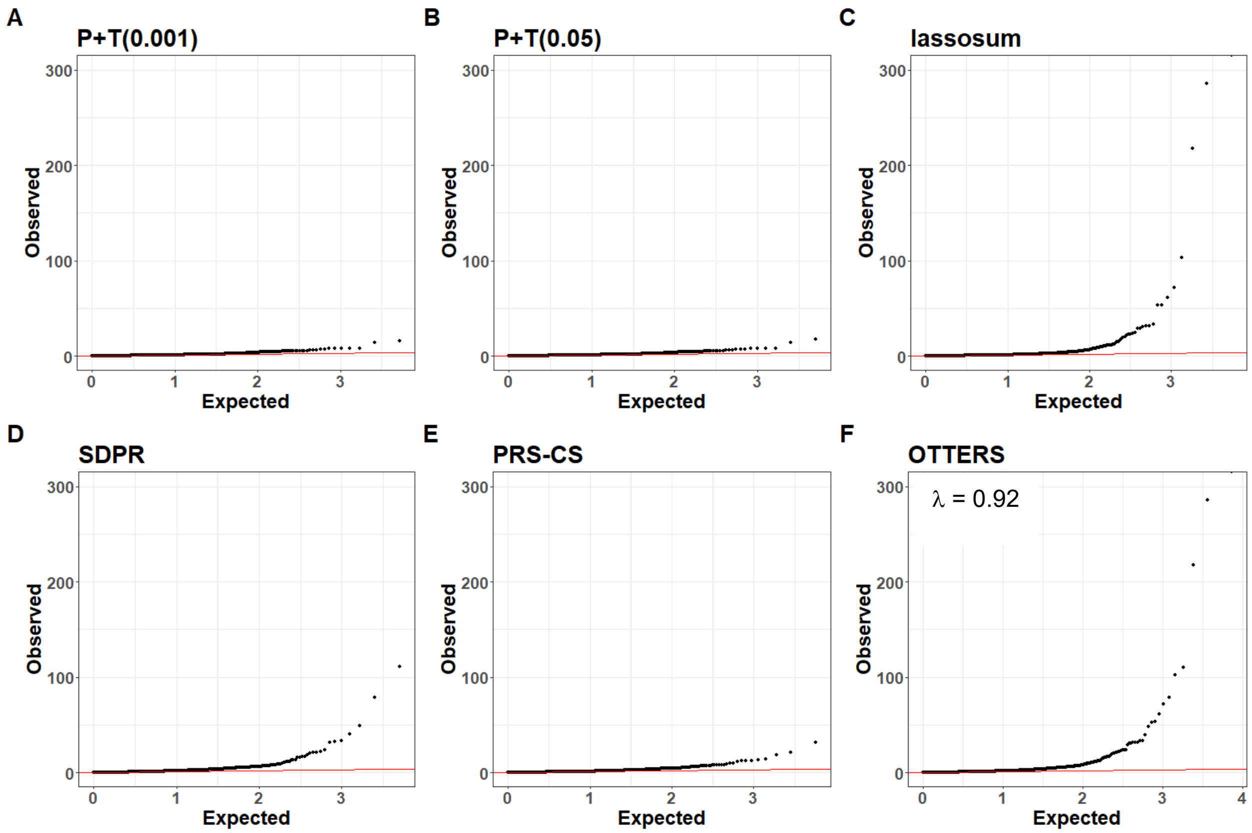


**Supplemental Figure 8.** Quantile-Quantile (QQ) plots of Blood TWAS p-values applied to Alzheimer's Disease and Related Dementias (ADRD) data of 5,149 genes by P+T(0.001) (A), 4,977 genes by P+T(0.05) (B), 5,406 genes by lassosum (C), 4,973 genes by SDPR (D), 5,672 genes by PRS-CS (E), and 7,241 genes by OTTERS (ACAT) (F). P-values are genomic-control corrected p-values generated from TWAS Z-score tests (two-sided). Only genes with valid models (Testing R > 0.1) are included within this plot.

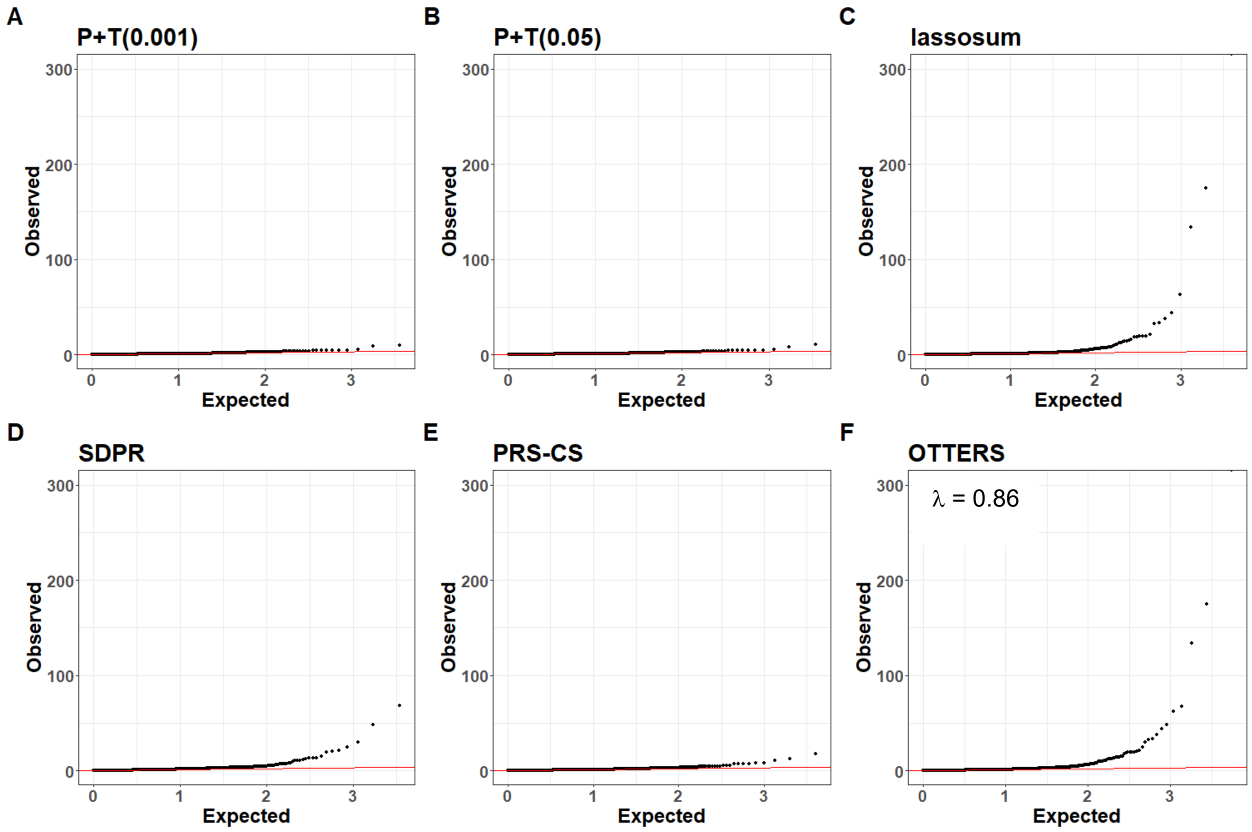


**Supplemental Figure 9.** Quantile-Quantile (QQ) plots of Blood TWAS p-values applied to Alzheimer's Disease and Related Dementias (ADRD) data of 3,579 genes by P+T(0.001) (A), 3,440 genes by P+T(0.05) (B), 3,961 genes by lassosum (C), 3,422 genes by SDPR (D), 3,984 genes by PRS-CS (E), and 5,489 genes by OTTERS (ACAT) (F). P-values are genomic-control corrected p-values generated from TWAS Z-score tests (two-sided) using only gene models with concordant effects across all modeling approaches. Only genes with valid models (Testing R > 0.1) are included within this plot.
